# Comparative evaluation of HIV testing interventions for men who have sex with men in the Netherlands: insights for a low-incidence setting

**DOI:** 10.64898/2026.03.16.26348499

**Authors:** Alexandra Teslya, Jacob Aiden Roberts, Janneke Cornelia Maria Heijne, Maarten Franciscus Schim van der Loeff, Ard van Sighem, Axel Jeremias Schmidt, Kai J Jonas, Mirjam E Kretzschmar, Ganna Rozhnova

## Abstract

**Background:** Although the number of new HIV diagnoses among men who have sex with men (MSM) in the Netherlands has declined considerably, the recent plateau suggests ongoing transmission. In 2024, 29% of new diagnoses among MSM were in a late HIV stage, showing that the time between infection and diagnosis can still be substantially reduced. In low-incidence settings, infections introduced through immigration are increasingly important in sustaining transmission, highlighting the need to re-evaluate current testing guidelines. We assess targeted testing strategies among MSM in the Netherlands addressing these considerations.

**Methods:** We used an agent-based model of HIV transmission among MSM in the Netherlands, incorporating infections acquired domestically and abroad. For 2024 - 2040, we simulated testing interventions targeting different subgroups, including offering an HIV test to immigrants upon entry, increasing testing rates among MSM residing in the Netherlands, and combinations of these approaches.

**Results:** Offering HIV testing to immigrating MSM at the entry averted up to 94 (95-th % quantile interval, 95% QI -128 – 328) new infections over 15 years if at least 50% take the test. Increasing testing to every 7 months in the general MSM population achieved the largest reduction, with up to 508 (95% QI 292 – 900) infections averted. The same testing rate in MSM with more than 5 partners within the previous six months resulted in 340 (95% QI 132–592) infections averted. Combining testing at entry with 7-months testing among general resident MSM averted the most infections, 534 (95% QI 308 – 884).

**Conclusions:** Combination of offering HIV test to immigrating MSM at the entry with 7-month testing frequency in the general resident MSM population can substantially reduce HIV infections. The difference in impact between targeting general MSM and those with relatively high recent partner numbers suggests that criteria for being at risk of having HIV need to expand.

**Author summary:** While HIV transmission among MSM in the Netherlands has decreased substantially over the last decade, it is still ongoing. In 2024, 29% of new HIV diagnoses in MSM were in individuals in late-stage of HIV infection, suggesting that the time between HIV acquisition and diagnosis should be shortened further. Additionally, in a low-incidence setting such as MSM in the Netherlands, introduction of HIV infections through immigration becomes more important. We evaluated several HIV testing strategies for this context, considering both immigrating MSM and resident MSM. While offering HIV test at entry point can avert many HIV infections, increasing testing rate in resident MSM to on average every seven months can avert substantially more HIV infections. The greatest impact is achieved when these approaches are combined: targeting both immigrating MSM and those already living in the country. This combined strategy requires the fewest additional tests per infection averted. Importantly, our simulations show that there are MSM living with undiagnosed HIV who do not necessarily meet the traditional criteria for being at risk. Improved testing strategies can help reach these individuals earlier, benefiting both public and their personal health.

## 2 Introduction

There has been a substantial progress in stemming the HIV epidemic among men who have sex with men (MSM) in the Netherlands. Both biomedical breakthroughs, such as the antiretroviral therapy (ART) and pre-exposure prophylaxis (PrEP), and decades of continued investment in treatment and prevention programmes have led to a sustained reduction in the rate of new HIV infections. By the end of 2023, levels of care engagement and retention within the MSM population exceeded the UNAIDS 95-95-95 targets set for 2030. It is estimated that over 96% of MSM living with HIV are diagnosed, 96% of those diagnosed receive ART, and 97% of those on ART have achieved viral suppression [42]. Additionally, the nationwide PrEP programme, rolled out in 2019, is credited with contributing to further declines in onward transmission [23, 45]. Despite these gains, MSM remain a key population, accounting for more than half of new HIV diagnoses each year [34, 35, 36, 37, 38, 39, 40, 41, 42]. The sustained rate of new HIV diagnoses in this population indicates ongoing HIV transmission.

The continued transmission may be attributed to a combination of factors, one of which is testing frequency. Although MSM at high risk for HIV acquisition are actively being engaged in HIV care and prevention services and offered frequent testing and other preventive measures, the recent report by Stichting HIV monitoring foundation (SHM) shows that 30% of new diagnoses were in individuals with a late stage of HIV infection, indicating opportunities for earlier detection [42]. In the modeling study assessing effectiveness of increased HIV testing among MSM in the Netherlands [22] Reitsema et al. showed that increasing testing uptake can substantially reduce new infections but is likely to be efficient only when targeted at MSM with high number of partners (defined as greater than 10 partners in the preceding 6 months). However, narrowing the focus group of the intervention results in fewer infections averted overall than for a broader testing target groups. This finding is in agreements with the conclusions of a scoping review by Brunner et al. [2], which reasoned that in the Western high-income countries with concentrated epidemics, HIV/STI prevention measures which target individuals with high number of partners are the most efficient. A USA-based study [44] showed that as the HIV prevalence decreases, the efficiency of routine screening programmes decreases (while still remaining worthwhile). Overall, evidence indicates that semiannual testing [11] and large-scale screening of populations at high risk for HIV acquisition [6] can result in faster HIV diagnoses and higher HIV infections averted, however, efficiency is context-dependent and tends to be higher when delivered in clinical settings and targeting individuals with high risk of HIV acquisition.

As the HIV epidemic in MSM in the Netherlands in recent years was characterized by a steady low incidence of new HIV infections, testing strategies must be refined to identify cases that current screening advisories do not capture. Screening guidelines often highlight individuals with recent sexually transmitted infections (STIs) or a large number of casual sexual partners. However, in populations with low HIV incidence such as MSM in the Netherlands, this approach may be less efficient for identifying HIV infections soon after acquisition. Combined, these considerations suggest that the exploration of testing approaches that extend beyond currently targeted groups and consider pathways of transmission from outside the local context may be both impactful and effective in a low-incidence setting where further progress requires more refined strategies.

In addition to HIV infections acquired through transmission networks among residents, a fraction of new HIV cases in the Netherlands appears to be individuals who acquired HIV abroad. This hypothesis is supported by recent mathematical modelling studies [20, 4, 23], which highlight the role of infections acquired abroad in sustaining transmission in low-incidence settings. For example, Teslya et al. [30] showed that even under intensified testing policies capable of increasing the detection of early infections, onward transmission is expected to persist, largely due to imported HIV infections and delayed diagnoses in individuals not reached by frequent screening. At the same time, studies have found that a substantial fraction of individuals at risk of having HIV, particularly those with an immigration background, have never been tested for HIV [10, 32]. Migrants entering the MSM population may arrive with an undiagnosed HIV infection, face barriers to accessing care, and experience delays in testing. To address these gaps, testing strategies must consider health realities of individuals with migration status. One-time HIV testing of incoming MSM immigrants could provide a relatively low-cost opportunity to detect infections early and reduce onward transmission, yet its potential impact has not been evaluated for the Dutch setting. In this work we aim to assess and compare the projected impact and efficiency of HIV testing interventions targeting incoming MSM as well as resident MSM in low-incidence setting similar to modern Netherlands. We extend a previously developed agent-based model [30] and evaluate testing interventions focusing on three strategies: (1) offering one-time HIV testing to incoming immigrants at entry point, (2) ensuring regular testing at a minimum rate among MSM with more than five or more than ten partners in the past six months, and (3) ensuring regular testing at a minimum testing rate across the general MSM population. We also assess combined strategies that target both infections arriving via immigration and those arising from local network dynamics, specifically by pairing intervention (1) with intervention (2), and separately with intervention (3). By comparing the projected impact and efficiency, we aim to derive actionable guidance for public health professionals to further decreasing onward HIV transmission among MSM in the Netherlands.

## 3 Methods

### 3.1 Model

Our modeling framework is an extension of a previously developed agent-based model of HIV transmission among MSM tailored to the questions of this study [30]. We updated the framework by implementing an explicit, risk-stratified testing module (with testing distributions varying by individual risk score), enabling analyses that were not supported by the original model. The model includes three key processes shaping HIV dynamics in this population: demography, dynamic sexual network, and HIV epidemiological process. Demographic processes include population entrance, exit, and ageing. The sexual network dynamics capture the formation and dissolution of two types of partnerships: long-term monogamous (steady) partnerships and short-term, possibly concurrent (non-steady) partnerships. Processes determining HIV epidemiology are HIV transmission, natural history, linkage to care and treatment, and PrEP programme. The full description of the model can be found in the Supplementary Material 1. Below, we give more details of each component of the model.

### 3.1.1 Demographic dynamics

We simulate the dynamics of a population of individuals aged between 15 and 75 (ages of sexual activity). Individuals enter, age, and leave the population. They exit the population either due to the background age-dependent mortality, AIDS-related mortality, or upon reaching the age of 75, which is used as a proxy for cessation of sexual activity. Mortality rates are calculated from publicly available data for the Netherlands [17]. The overall entrance rate is calculated to maintain a population size of 25,000 individuals. Model outcomes are scaled to a population of 200,000, the estimated number of MSM in the Netherlands [12]. The entrance to the population mimics reaching the age of sexual debut, thus, the age of individuals upon entrance follows the distribution of ages at sexual debut as described by the European MSM Internet Survey data from 2017 (The European MSM Internet survey 2017, EMIS-2017) [46].

#### Immigration

While most individuals who enter the population do not have HIV, we account for immigration by modeling a proportion of the newly entered individuals as having HIV, either diagnosed or undiagnosed. To estimate the annual number of incoming immigrants to the Netherlands, we have consulted immigration rates presented by Statistics Netherlands [18] over 2014–2019 and projected these rates for years 2020-2040 using linear regression. Based on the estimated 1% of the Netherlands population being MSM, we assumed the same proportion among incoming immigrants and projected the annual number of MSM arrivals accordingly. Over 2017-2023 SHM estimated that each year between 87% to 95% of individuals living with HIV who arrive in the Netherlands already aware of their HIV status have started ART. Due to estimated shorter time between HIV acquisition and diagnosis in MSM compared with the general population, and the robust state of cascade of care, we model that 95% of incoming MSM living with HIV and aware of their status, have a suppressed viral load. We model that all individuals who know their HIV status engage with HIV care services and proceed through the care stages at the same rate as the residents. The entrance rate of individuals with undiagnosed HIV was obtained through calibration of model outputs to the data (see Figure 3, Section 1.4.2 in Supplementary Material 1 for the details).

#### 3.2.1 Progression of HIV infection

We use a standard framework that divides infection progression into four stages: a relatively brief primary stage with high infectivity, a prolonged chronic stage with lower infectivity, an AIDS stage with increased infectivity and AIDS-related mortality, and a late AIDS stage marked by very high infectivity, absence of sexual activity and low expected survival.

#### 3.2.1 Care and prevention programmes

##### PrEP

In the Netherlands, a national PrEP programme that subsidized the cost of PrEP for eligible individuals was launched in 2019. Following its rollout, until 2024, the number of participants was capped by 8,500 individuals [23]. In the model, the programme was assumed to start in 2019 with zero participants, gradually increasing to a maximum of 10,000 individuals over 3 years, after which the number of individuals using PrEP remains constant. The upper bound for individuals who use PrEP in the model exceeds the programme’s capacity to account for individuals obtaining PrEP through other channels. The guidelines for the programme enrollment consist of criteria that identify individuals at increased risk of acquiring HIV. In the model, each individual has an intrinsic property, referred to as propensity to acquire casual partners which serves as a proxy for these criteria. Subsequently, individuals whose propensity to acquire casual partners exceeds a certain threshold are eligible to enroll. The threshold is a parameter determined through calibration.

##### HIV testing

The uptake rate depends on the recent number of non-steady partners [30]. Guided by Visser et al. [43], we stratify the population into four groups by the number of non-steady partners in the last six months: 0-2, 3-4, 5-10, and more than 10 partners. Individual testing frequencies are sampled from a distribution specific to each partner-number group, such that individuals who, on average, have fewer non-steady partners test less frequently than those with more non-steady partners. An additional testing category includes individuals enrolled in the PrEP programme. In the model, this group adheres to the programme’s testing guidelines: prior to January 2025, individuals who use PrEP test for HIV every three months [23], and thereafter every six months [16].

##### Cascade of care

Individuals diagnosed with HIV are immediately linked to care and, after an interval of time, initiate ART. This is followed by a period leading to viral suppression, after which the risk of virus transmission through anal intercourse is eliminated. The model allows for dropping out of care/treatment failure, and individuals are eligible to re-start the treatment or change to second line treatment.

### 3.2 Parametrization and calibration

To obtain model parameter values, we use data relevant to the MSM population in the Netherlands. Some parameters are directly computed from the available data, while the remaining parameters are estimated by calibrating model outputs to observed sexual network and epidemiological dynamics. To obtain parameters governing the sexual network structure and within-partnership behavior, we use data from the EMIS-2017 [46], the longitudinal Amsterdam Cohort Studies (ACS) [8], and the Amsterdam MSM Network Study [7, 13].

Some model parameters related to HIV transmission dynamics are informed by the literature [9, 24, 25, 43], while we obtain the remaining parameters by calibrating model outputs to the annual number of new HIV diagnoses, the time series of the care cascade and estimated annual distribution of time since infection at the time of diagnosis using national HIV surveillance data for MSM from 2017 to 2024 [35, 36, 37, 38, 39, 40, 41, 42].

Through the calibration of the HIV transmission process, we select 100 parameter sets that result in the closest fit of model outputs to observed data trends, based on the median simulated values. Details of the parameter estimation procedure along with emergent parameter values and their ranges are given in Table **7** and Figure **4** in Supplementary Material 1.

### 3.3 Simulation procedure

The model is calibrated through 2017–2023, in 2024–2025 runs forward without added interventions other than HIV care services and PrEP programme currently active in the Netherlands, and then interventions are applied from 2026 through 2040. Each intervention scenario is simulated using 100 parameter sets obtained through the calibration process, with 20 trajectories per parameter set, resulting in 2000 simulated trajectories per scenario.

### 3.4 Model output

In each scenario, we calculate the following outputs: annual number of new HIV infections, HIV diagnoses and HIV tests taken, and annual distributions of time since infection at the time of diagnosis in the intervention period. We also calculate the cumulative number of HIV infections averted, change in the cumulative number of new HIV diagnoses, and annual average number of additional HIV tests per 100,000 individuals over a 15-year intervention period relative to the baseline. The impact of interventions is estimated as the cumulative number of new HIV infections averted over 15-year period while efficiency of the interventions is given as an average number of additional HIV tests taken per HIV infection averted. We summarize the outputs by calculating medians for each parameter set, and then calculate the median and 95th quantile interval (95% QI) of the medians across 100 parameter sets. To compare all intervention scenarios and identify those performing optimally under specific constraints, we conducted a Pareto frontier analysis, evaluating each intervention by (i) the number of HIV tests per infection averted versus the cumulative infections averted, and (ii) the average annual number of HIV tests per 100,000 individuals versus the cumulative infections averted. In the figures the projections are presented in terms of medians with the full descriptive statistics of outputs is given in Table **1** in Supplementary Material 2.

### 3.5 Calibration

The projections of the model for years 2017-2023 align well with data trends on the annual number of new HIV diagnoses, the time since HIV acquisition at the time of diagnosis, and the evolution of the cascade of care in 2017-2023 (see Figure **4** in Supplementary Material 1).

By calibrating the model to HIV surveillance data we obtained the average annual inflow rate of new HIV infections from abroad, estimated at median 39 cases (95th QI 26–48) per year.

Since the available empirical estimates of testing frequency by risk group are relatively coarse and based on older data, we treated the testing rates as calibration parameters and, using the empirical values as a starting point, adjusted them so that the model reproduces the annual distribution of time since infection at diagnosis reported in the SHM surveillance reports. The grid of testing rates is shown in Table 1). The table lists the testing intervals obtained in the calibration process and used in the model (first column), their equivalent annual rates in tests per person per year (second column) and the probability of having been tested within one year (third column). The 3- and 6-month intervals (marked in Table 1) reflect PrEP programme recommendations and are applied only to individuals using PrEP (see note in Table 1).

**Table 1:** Conversion of testing intervals to annual testing rates and corresponding probabilities of testing within 1 year. The first column lists testing intervals obtained in the calibration process.

| Testing interval | Testing rate (per year) | Probability tested Within 1 Year |
| --- | --- | --- |
| Every 10 years | 0.10 | 0.10 |
| Every 5 years | 0.20 | 0.18 |
| Every 23 months | 0.52 | 0.41 |
| Every 7 months | 1.71 | 0.82 |
| Every 6 months* | 2.00 | 0.86 |
| Every 3 months* | 4.00 | 0.98 |

\* These intervals reflect testing recommendations from the PrEP programme.

Table 2 summarizes how individuals who do not use PrEP are distributed across the four main testing-frequency tiers. Each column in Table 2 corresponds to a partner-number group (defined by the number of non-steady partners in the previous six months), and the column entries give the probability that a person in that partner-number group falls into each testing-interval category. Partner-number groups are:

- Partner-number group 1: 0–2 non-steady partners (lowest number of partners);
- Partner-number group 2: 3–4 non-steady partners;
- Partner-number group 3: 5–10 non-steady partners;
- Partner-number group 4: >10 non-steady partners (highest number of partners).

**Table 2:** Proportions of MSM per HIV testing interval by partner-number group. Partner-number group 1 denotes individuals with 0–2 non-steady partners in the previous 6 months, 2 - individuals with 3–4 partners, 3 - 5–10 partners, and 4 - more than 10 partners. Individuals which belong to a higher partner-number group test more frequently on average. Note: these distributions over testing intervals apply to individuals who are not using PrEP.

| Testing Interval | Partner-number group 1 | Partner-number group 2 | Partner-number group 3 | Partner-number group 4 |
| --- | --- | --- | --- | --- |
| Every 10 years | 0.15 | 0.30 | 0.33 | 0.14 |
| Every 5 years | 0.50 | 0.26 | 0.15 | 0.26 |
| Every 23 months | 0.23 | 0.28 | 0.33 | 0.33 |
| Every 7 months | 0.12 | 0.16 | 0.19 | 0.27 |

Columns in Table 2 sum to one, so that for each partner-number group the table gives a probability distribution over the discrete testing-frequency tiers. (Individuals who use PrEP are handled separately and therefore not represented in Table 2.)

In the baseline (no-intervention) scenario, each individual who is not using PrEP is assigned a testing interval by sampling from the distribution for their partner-number group (Table 2); individuals who use PrEP are assigned the 3- or 6-month interval, depending on the calendar time within simulation.

For computational tractability, we define three intervention testing target tiers (corresponding to rows 2, 3, and 4 of Table 1): (1) testing once every 5 years; (2) testing once every 23 month; (3) testing once every 7 months. These three tiers were selected since they correspond to the calibrated testing frequencies which we subsequently deem as realistic policy targets (see Table 1).

### 3.6 Scenarios

The scenarios under consideration fall into two categories: baseline and intervention. In the baseline scenario, no additional interventions are deployed, and the parameters of the ongoing HIV care and prevention programmes remain unchanged from the values used in the calibration period. Table 3 lists all scenarios with target populations and parameter values.

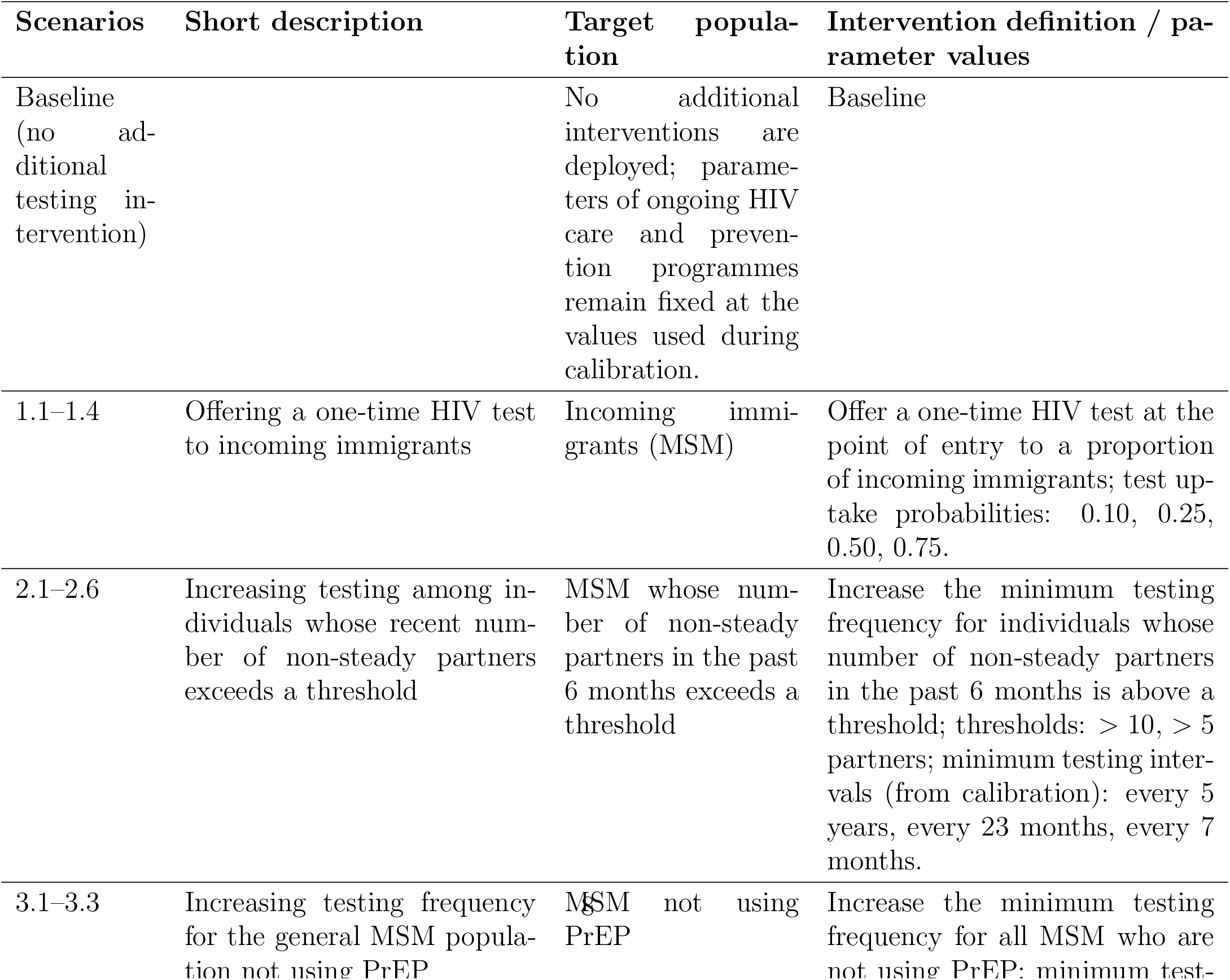

| Scenarios | Short description | Target population | Intervention definition / parameter values |
| --- | --- | --- | --- |
| Baseline<br>(no additional testing intervention) |  | No additional interventions are deployed; parameters of ongoing HIV care and prevention programmes remain fixed at the values used during calibration. | Baseline |
| 1.1–1.4 | Offering a one-time HIV test to incoming immigrants | Incoming immigrants (MSM) | Offer a one-time HIV test at the point of entry to a proportion of incoming immigrants; test uptake probabilities: 0.10, 0.25, 0.50, 0.75. |
| 2.1–2.6 | Increasing testing among individuals whose recent number of non-steady partners exceeds a threshold | MSM whose number of non-steady partners in the past 6 months exceeds a threshold | Increase the minimum testing frequency for individuals whose number of non-steady partners in the past 6 months is above a threshold; thresholds: > 10, > 5 partners; minimum testing intervals (from calibration): every 5 years, every 23 months, every 7 months. |
| 3.1–3.3 | Increasing testing frequency for the general MSM population not using PrEP | MSM not using PrEP | Increase the minimum testing frequency for all MSM who are not using PrEP; minimum test- |

When an intervention enforces a testing interval (e.g., once every 23 months), any individual whose assigned testing interval is longer than that minimum is reassigned to the minimum interval. Depending on intervention, this applies either to all individuals (population-wide increase), or only to individuals in specific partner-number groups.

## 4 Results

### 4.1 Baseline projections

The model projects a median of 1,178 (95th QI 840–1,596) new HIV transmissions within the Netherlands over the period 2026–2040. During the same period, 1,724 (95th QI 1,344–2,148) new HIV diagnoses are expected, including 164 (95th QI 120–228) in individuals who acquired HIV within the past 3 months, 120 (95th QI 88–160) in those who acquired HIV 3–6 months earlier, and 228 (95th QI 160–300) in those who acquired HIV 6–12 months earlier. The projection that cumulative new HIV diagnoses exceed cumulative new HIV infections indicates that a proportion of individuals diagnosed in 2026—2040 either acquired HIV before 2026 or acquired it abroad. On average, 60,903 HIV tests per 100,000 individuals (95-th QI 53,684–68,857) are taken yearly.

### 4.2 Impact of interventions

We assess the impact of interventions by analyzing the cumulative number of new HIV infections averted compared to the baseline scenario over 15-year time horizon (Figure 1). Overall, greater impact is expected from combination interventions, although their effect is comparable to the additive contributions of the single-group interventions that comprise them.

**Figure 1:**
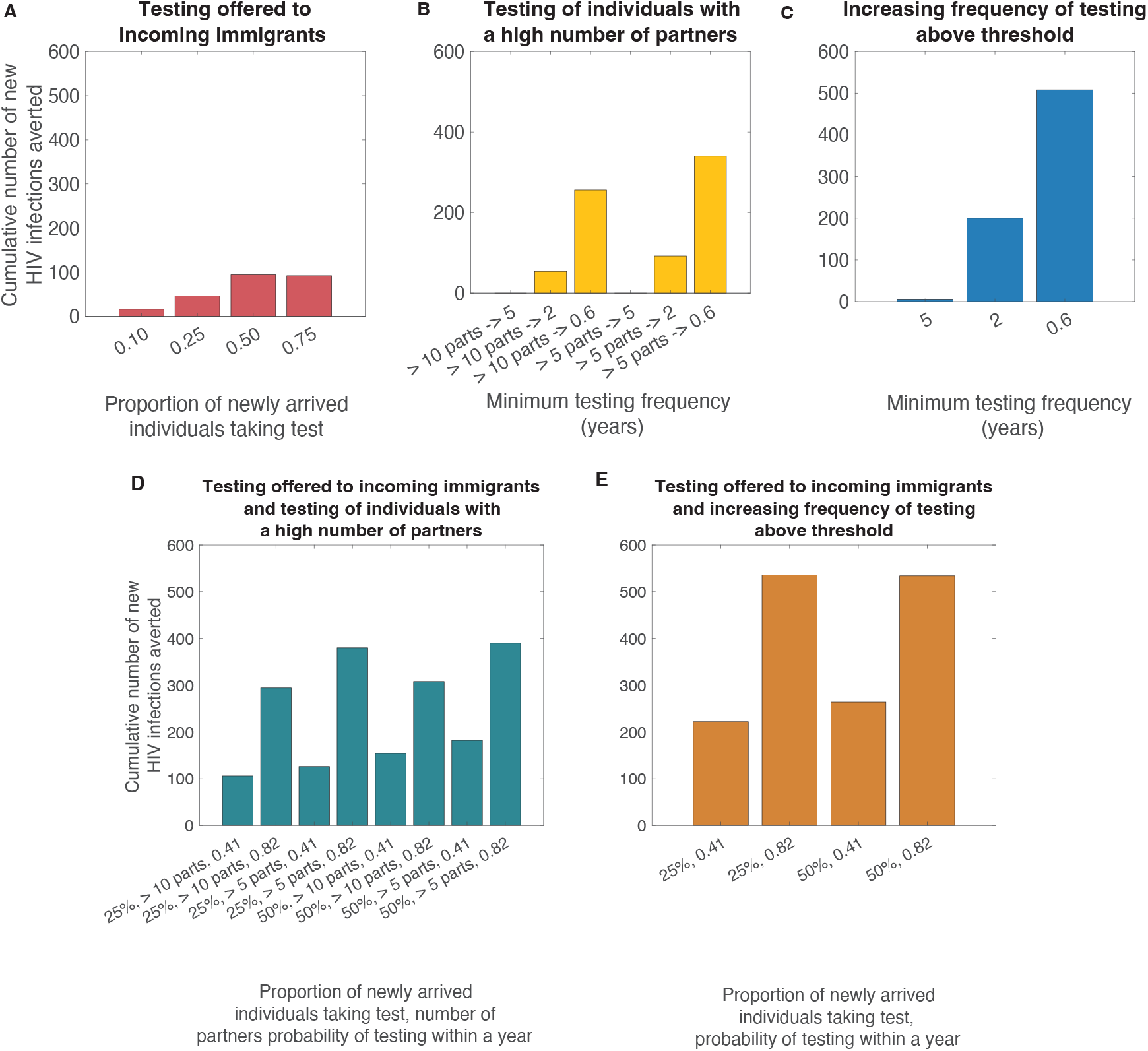
Cumulative number of new HIV infections averted. **A** Offering a one-time HIV test to incoming immigrants. **B** Increased testing of individuals with the number of non-steady partners in the last 6 months exceeding threshold. **C** Increasing probability to test within 1 year. **D** One-time testing of individuals immigrating to the Netherlands combined with increased testing of individuals with the number of non-steady partners in the last 6 months exceeding threshold. **E** One-time testing of individuals immigrating to the Netherlands combined with increasing probability to test within 1 year.

Among the interventions which consider a single group (interventions 1 through 3), the largest number of new HIV infections averted is achieved by increasing testing frequency in the general population of MSM to be on average every 7 months, resulting in 508 (95% QI: 268–740, Figure 1 **A–C**) infections averted. The next best outcome is expected for intervention which targets individuals with more than 5 partners in the last 6 months and achieves a testing frequency of every 7 months in this group, with 340 (95% QI: 132–592) new HIV infections averted. Finally, if individuals with more than 10 partners in the past six months test at least every 7 months, the number of averted HIV infections is 256 (95% QI: 68–464). The intervention where HIV testing is offered to incoming immigrants at the entry point is projected to avert at most 94 (95% QI: -128–328) new HIV infections (Figure 1 **A**). Interestingly, this occurs when 50% of offered tests are taken, the outcome which is not improved at 75% test uptake with 92 (95% QI: -60–344) new HIV infections averted, respectively.

The greatest impact is observed for the intervention which combines offering one-time test to incoming immigrants with increasing the testing rate in the general population of MSM to be on average every 7 months, corresponding to an annual testing probability of 0.82. For such interventions, the cumulative number of HIV infections averted over the 15-year period is equal to 536 (95 % QI: 312–792) for testing probability for incoming immigrants of 0.25 and to 534 (95 % QI: 308–884) when the testing probability is equal to 0.5 (Figure 1**E**). It is noteworthy, that while these results are an improvement on the next best intervention, which increases the testing rate in the general population of MSM to every 7 months (500 (95 % QI: 268–740), (Figure 1**C**), the difference is rather small. The next best outcome of 390 (95% CI: 180–780) averted HIV infections is expected for the intervention which combines offering test to incoming immigrants which is accepted with rate 0.5 with increasing testing threshold in individuals who had more than 5 partners in the previous 6 months to every 7 months (Figure 1**D**). If the strategy results in only 25% of incoming immigrants taking the test, the projected gain is comparable at 380 (95% QI: 148–672) HIV infections averted.

Time series analysis of annual number of new HIV infections under different scenarios (Figure **2**, in Supplementary Material 2) shows that for interventions with sizeable cumulative number of new HIV infections averted (all interventions whose component involves resident MSM) the incidence of new HIV infections decreases drastically compared to the baseline in the first two years following intervention launch, thereafter settling on a plateau. In scenarios with significant incidence reductions, the annual diagnosis time series exhibits an increase in the first few years after the intervention start followed by a decline below baseline in later years, a pattern consistent with earlier detection preceding reduced onward transmission (see Figure 3, Section 2.1.2, Supplementary Material 2).

### 4.3 Efficiency of interventions

We assessed efficiency of each intervention in terms of the average annual number of additional HIV tests performed per 100,000 individuals per HIV infection averted (Figure 2).

**Figure 2:**
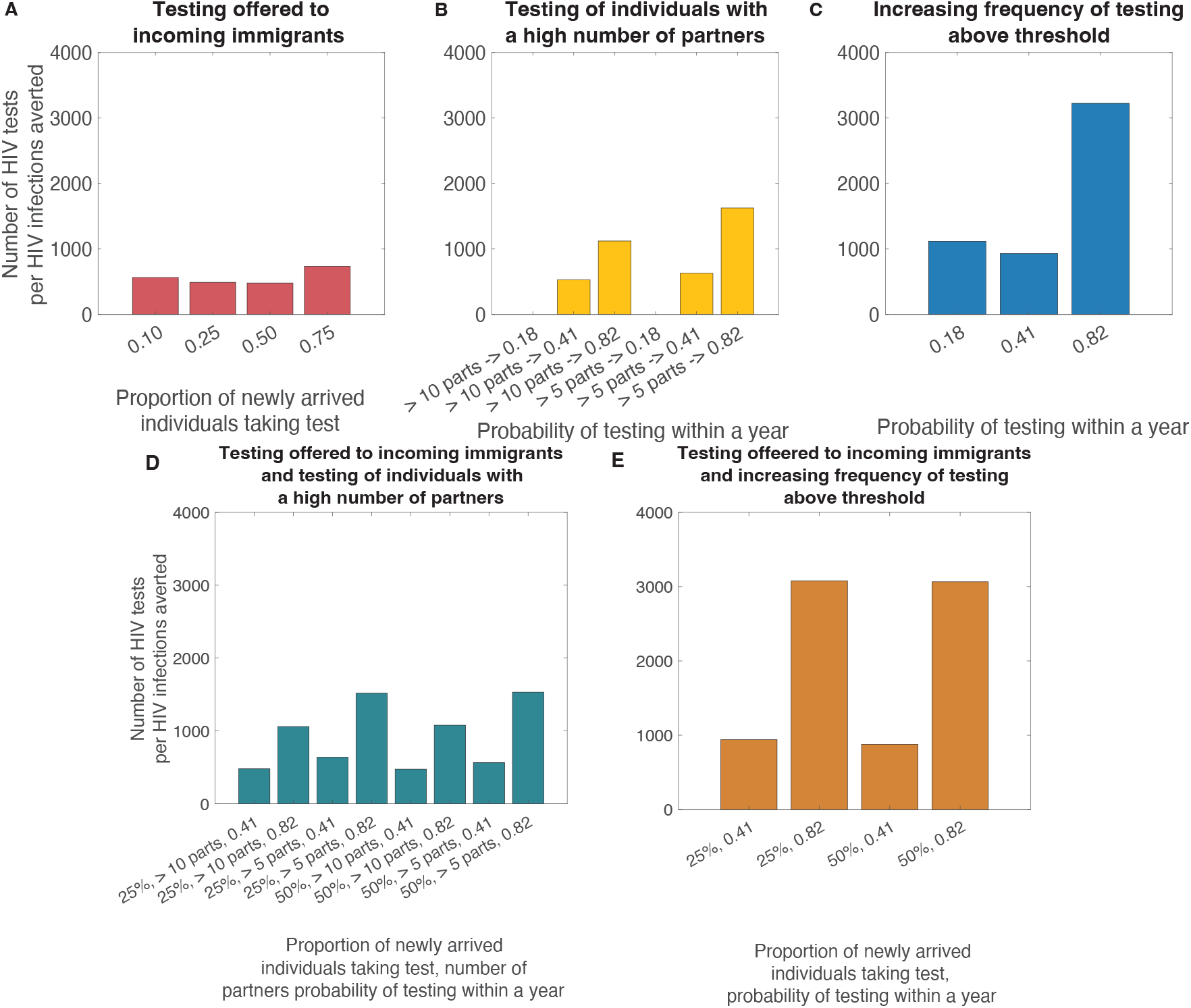
Efficiency analysis for HIV infection averted. **A** Offering a one-time HIV test to incoming immigrants. **B** Increased testing of individuals with number of non-steady partners in the last 6 months exceeding threshold. **C** Increasing probability to test within 1 year. **D** One-time testing of individuals immigrating to the Netherlands combined with increased testing of individuals with number of non-steady partners in the last 6 months exceeding threshold. **E** One-time testing of individuals immigrating to the Netherlands combined with increasing probability to test within 1 year. Efficiency is shown only for interventions with positive median of the number of HIV infections averted.

**Figure 3:**
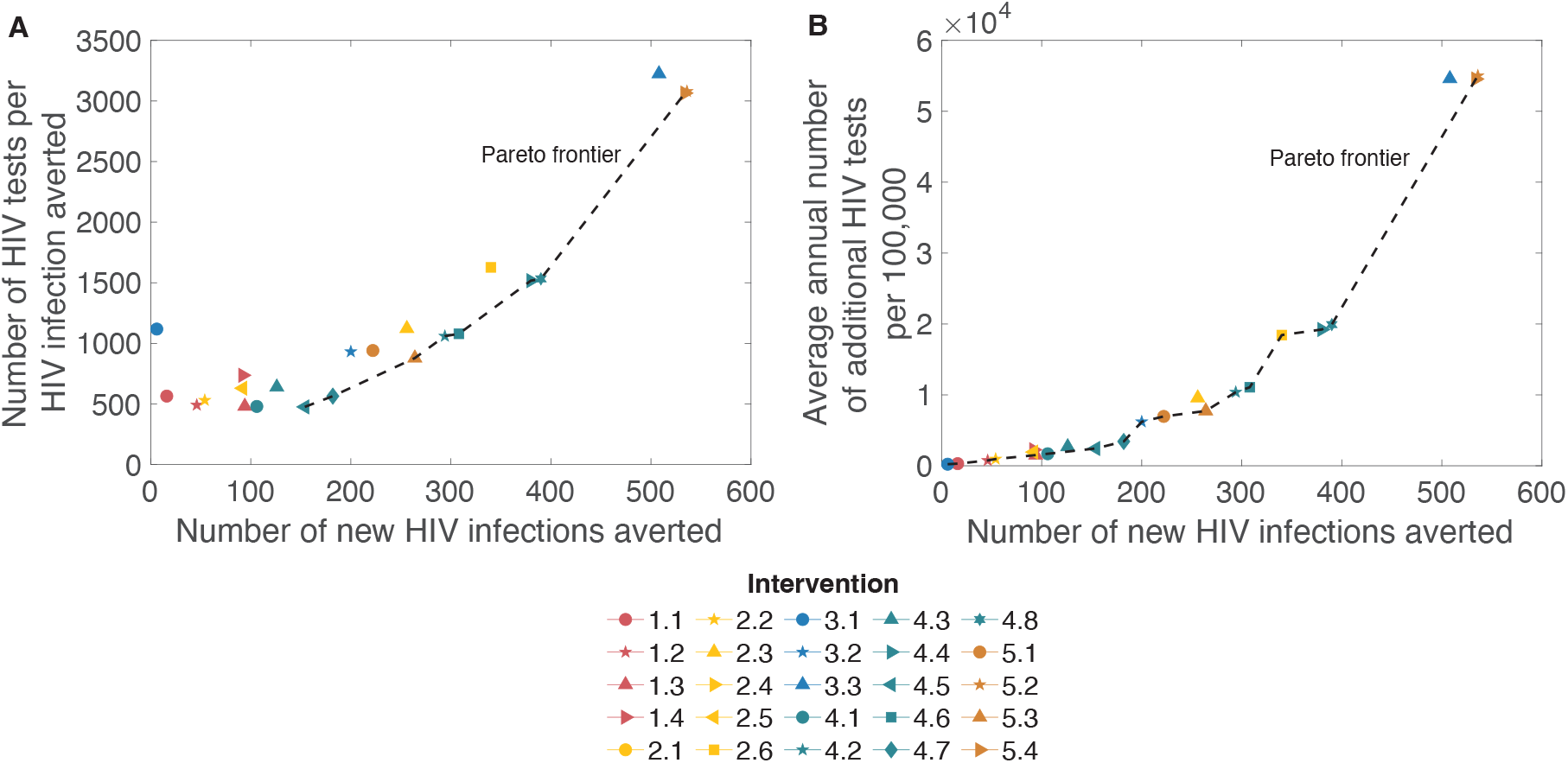
Comparison of interventions in terms of impact and efficiency. **A** shows average number of additional HIV tests per HIV infection averted vs cumulative number of new HIV infections averted. **B** shows average annual number of additional HIV tests per 100,000 individuals vs cumulative number of new HIV infections averted. Punctured line shows Pareto frontier calculated over median values for each considered scenario. Only interventions with positive median of the number of HIV infections averted are shown.

Overall, we observe that the efficiency of combination interventions (Interventions 4 and 5, Figure 2**D** and **E**) is comparable to the interventions targeting a single group within residents of the Netherlands (Interventions 2 and 3, Figure 2**B** and **C**).

For the intervention where testing is offered incoming immigrants, we observe that for testing uptake between 0.10 and 0.50, increasing testing rate leads to increased efficiency. The increase does not continue when the proportion of incoming immigrants taking the test increases to 0.75, as the number of HIV infections averted remains similar to that for 0.5 testing uptake, but the number of individuals who take the test increases significantly. For interventions where individuals with a high number of partners test more frequently, as the testing rate grows the efficiency of intervention declines. For a testing frequency of every 7 months increase in the target group size (from individuals with more than 10 partners in the last 6 months to individuals with more than 5 partners over the same period), there is a modest decrease in efficiency, from 1,122 (95 % CI: 665–3,961) to 1,627 (95 % CI: 997–3932) HIV tests per HIV infection averted. In contrast, for the intervention achieving the general testing frequency of every 7 months, the efficiency of the intervention is projected to be 3,305 (95 % CI: 2,486–5,549) HIV tests per HIV infection averted. Thus, while efficiency decreases with increasing the scope of the targeted population, the overall impact in terms of HIV infections averted increases.

Finally, we compare the interventions in terms of their impact versus efficiency (Figure 3). We have considered only intervention scenarios that are projected to result in positive number new HIV infections averted. We observe that in the intervention where immigrating MSM take a test, on the range of 0.1–0.50 the efficiency of the intervention increases as the probability of taking the test increases, and the number of HIV infections averted increases, respectively. However, uptake of tests equal to 0.75 does not result in additional HIV infections averted and sharply decreases the efficiency. Overall, the impact of this intervention is significantly smaller than the impact projected for the remaining two single-group interventions. Generally, we observe that the most impactful intervention is increasing the testing frequency in the general population of MSM to every seven months, resulting in between 534 to 536 new HIV infections averted over 2026– 2040 with an efficiency of 3,064 and 3,079 HIV tests per HIV infection averted, for combination interventions 5.2 and 5.4. A comparable outcome of 500 new HIV infections averted with 3,305 HIV tests per HIV infection averted, intervention 3.3. Interventions which includes the component where all individuals with more than 5 partners within the last six months test every seven months result in the second best number of HIV infections averted and a worse efficiency with 340–390 HIV infections averted at the cost of 1,517–1,626 HIV tests per infection averted. To highlight best performing interventions we have calculated Pareto frontier for both analyses shown on Figure 3. With respect to optimizing number of additional HIV tests per HIV infection averted (Figure 3 **A**), the frontier consists only of combination interventions with requirement for either probability of taking of test within year being 0.82 or that at least 50% of incoming MSM take the test on entry. We observe that the frontier exhibits a distinct turning point: for strategies achieving up to 400 infections averted, additional impact can be obtained with comparatively modest increases in testing, while interventions which result in more than 500 infections averted the marginal testing requirement grows faster. On the other hand, when considering the Pareto frontier on Figure 3 **B** capturing annual number of additional HIV tests per 100,000 individuals vs cumulative number of new HIV infections averted, we observe that almost all interventions are Pareto dominating. We observe that the Pareto frontier curve can be roughly divided into three regions characterized by different growth in the number of tests per increment in additional infections averted: up to 200 infections averted, between 200 and 400, and more than 400, such that the growth in each region is higher than in the preceding, indicating fast growing marginal test requirement.

A full description of the projections in terms of cumulative number of new HIV infections averted, number of additional HIV tests per HIV infection averted, and annual number of additional HIV tests taken per 100,000 per year, all reported as medians with 95% quantile ranges, is given in Table **1** in Supplementary Material 2.

### 5 Discussion

We used an agent-based model of HIV transmission dynamics among MSM in the Netherlands to evaluate the impact and efficiency of testing interventions targeting different population groups. We specifically aimed to compare interventions targeting MSM newly immigrating to the Netherlands with those targeting MSM residing in the Netherlands. For a low-incidence epidemic, such as currently observed among MSM in the Netherlands, HIV infections already present at the time of immigration increase in importance for onward HIV transmission. We focused on this population by considering an intervention in which individuals immigrating to the Netherlands are offered HIV-related information and an optional HIV test at the point of entry. We compared the impact and efficiency of this intervention with those of testing strategies targeting resident MSM who have had recent high numbers of non-steady partners or those who test less frequently than the minimum threshold. We also considered combined interventions targeting both individuals who acquired HIV abroad at the point of entry and individuals already residing in the Netherlands.

Our study projects that offering an HIV test to incoming individuals at the point of entry could avert up to 92 new infections in MSM population over 15 years if at least 50% of those offered an HIV test accept it. A comparable outcome could be achieved if individuals with more than five partners in the previous six months increased their testing frequency to every 23 months. The efficiency of both approaches was similar, requiring a median of 480 and 630 tests per infection averted, respectively. Further increases in test uptake among incoming MSM produced negligible additional gains while reducing efficiency. The interventions where an HIV test was offered to incoming MSM with an uptake of 50% or 75% were projected to have the highest efficiency of all considered interventions, whether single-group or combined, with 491 and 480 additional HIV tests per infection averted, respectively. Among other single-group testing interventions, the largest impact was achieved when the general MSM population tested, on average, every seven months. However, this intervention had a significantly decreased efficiency, with 3,224 additional HIV tests per HIV infection averted.

In terms of impact, combination interventions were projected to take the lead; however, the total number of HIV infections averted did not exceed the sum of their component single-group interventions. In fact, when combined interventions included subgroups testing as frequently as every seven months, they averted fewer new HIV infections than the sum of their individual components. However, in terms of efficiency, as indicated by the number of additional HIV tests per HIV infection averted, combining the offering of HIV tests to newly arriving MSM with an intervention aimed at increasing the testing rate of the subpopulation of resident MSM almost always resulted in an efficiency that was higher than if the resident-targeting intervention had been deployed on its own. Moreover, this result becomes more noticeable as the testing rate for the resident subpopulation increases.

Depending on the available constraints, our analysis suggests several scenarios that may be more favourable. When the intervention duration and total budget are fixed, almost all considered scenarios can be suitable, with the exact choice depending on resource limit. Conversely, if the intervention runs indefinitely and the cost per infection averted is the primary constraint, our efficiency analysis suggests that combined intervention, targeting both immigrating and resident MSM, should be considered.

Overall, the MSM population in the Netherlands represents a low-incidence setting for HIV transmission [29]. In such contexts, PrEP emerges as one of the most considered interventions [3, 23, 45], yet testing uptake continues to play a crucial role. Testing interventions have already contributed to substantial reductions in HIV incidence, and further increases could yield additional, cost-effective gains [3]. However, as incidence declines, the cost-effectiveness of broad category-based programmes diminishes, and targeted strategies become increasingly important [33]. In our previous work [30], we showed that while increasing diagnosis rates among MSM with early HIV infection, combined with immediate ART initiation, could avert a significant number of new HIV infections, under such an intervention, a significant proportion of individuals living with HIV are missed. Reitsema et al. [21] demonstrated that testing recommendations based on recent partner numbers could prevent a significantly larger number of new HIV infections. While this study study focused solely on interventions increasing testing uptake, Reitsema et al. [23] demonstrated that increased testing among PrEP participants provides additional, though diminishing, benefits as the programme coverage grows. Combined, the results of this and previous studies indicate that to further reduce onward HIV transmission among MSM in a low-incidence setting, combination interventions should be considered, targeted at both the resident population and the newly arriving individuals [27].

While our model reproduces recent Dutch HIV surveillance trends, several simplifications may have influenced quantitative projections. In the model, HIV infections acquired outside the Netherlands were represented exclusively as infections entering exclusively through immigration. In reality, some infections may occur among Dutch residents during short-term travel abroad. We note that offering information and an HIV test in such a scenario would be logistically and financially more challenging. Consequently, our efficiency estimates, expressed as tests per infection averted, may be optimistic. To calculate the number of tests taken in the intervention when HIV testing is offered to incoming immigrants, we used the total estimate of immigrating MSM, rather than the general population. Thus, our estimates of the effectiveness of this intervention may have been overly optimistic.

Another simplification concerns behavioral and structural differences between resident and migrant MSM. The model assumes identical testing uptake, care linkage, and partnership formation patterns across these groups. However, studies from the Netherlands and Europe indicate that migrant MSM face greater barriers to HIV prevention and care than non-migrant MSM, and that a substantial proportion have never tested for HIV or do not test regularly, contributing to a higher risk of late diagnosis [28, 1]. Moreover, substantial heterogeneity in this respect exists across countries of origin [28]. Other aspects of the HIV care continuum, due to factors such as stigma, language barriers, and social connectedness, also vary between the Dutch-born residents and individuals with a migratory background [15]. Finally, a study by Santoso et al. has demonstrated [26] that HIV prevalence in the migrant population tends to be higher than in the native-born population, underscoring the additional HIV burden this subpopulation may be facing. Accounting for these differences in future work will be critical for refining projections of the impact of interventions that target immigrating MSM.

Recent study highlighted the importance of clear protocols and skilled staff for the effective delivery of entry-point HIV testing interventions [14]. While our study evaluates this intervention assuming effective implementation, a real-world rollout must be ethically grounded and sensitive to cultural differences that shape acceptability and trust. Testing should be strictly voluntary (optin), with clear multilingual consent, robust confidentiality and data protection, full separation from immigration or policing, and immediate, non-discriminatory linkage to care co-designed with key stakeholders [19, 5, 31]. Recent evidence has shown feasibility and acceptability of integrating HIV testing into migrant tuberculosis screening in the Netherlands, suggesting encouraging prospects for such an intervention [14].

In our analysis, single-group interventions that achieved testing rate of every seven months in a subgroup of the population averted 340–508. When comparing interventions that resulted in increased testing rates among MSM with more than five partners versus those that targeted the general MSM population, the latter produced markedly greater reductions in transmission, indicating that conventional behavioural criteria for testing may not be the most impactful and fail to reach individuals who fall outside these definitions. The number of recent partners may identify individuals who have recently acquired infection, but not those with a long-established infection. According to SHM [42], in 2023, 31% of MSM diagnosed with HIV were diagnosed in advanced stage of HIV infection characterized by CD4 counts below 350 cells/mm^3^ or having AIDS. Thus, targeting individuals who rarely or never test may be more effective. Leenen et al.[10] showed that MSM who never test in less urbanized areas tend to have low perceived severity of HIV infection and themselves at lower risk for acquisition, and have fewer gay friends and weaker social ties, while urban MSM who rarely test are often younger, bisexual-identifying, have fewer partners, and report no recent condomless anal intercourse. Together, these findings suggest that increasing testing uptake will require approaches that differ from conventional risk-based strategies and that these approaches may need to be tailored to local context.

Our results have shown that, in the current low-incidence HIV epidemic among MSM in the Netherlands, the most effective way to further reduce onward transmission is to combine raising the minimum testing rate across subgroups of the resident population to at least every 23 months and complement this with HIV test offer at entry for newly arriving MSM. The latter, in addition to its preventive effect, also facilitates faster linkage to care for individuals for whom engaging with the healthcare system may be slower and more challenging than for Dutch-born residents [15]. In an increasingly globalized world, migration will continue to shape the HIV epidemic, and testing policies designed primarily around resident populations may fail to recognise that migrants require distinct approaches and dedicated consideration. In this context, our analyses illustrate how entry testing could complement resident-focused strategies and highlight the need to consider how testing and linkage services are organised for migrants, alongside resident MSM, rather than primarily for those already well integrated into existing services.

To support such strategies, future modelling work should represent migrant-related epidemiological processes more explicitly, including behavioural heterogeneities in mixing patterns, condom use, testing frequency, ART adherence, and PrEP access. A more detailed representation of these differences may reveal greater potential for testing interventions targeting immigrating MSM, but such refinement will require dedicated data collection and parameterization.

## Supporting information

Additional analyses

## Data Availability

All data produced are available online at https://github.com/aiteslya/HIV\_MSM\_Testing\_AB.

## 6 Funding declaration

This work was supported by Aidsfonds (grant P-53902; awarded to GR, https://aidsfonds.nl/) and by the VERDI project (grant 101045989 awarded to GR, https://verdiproject.org/the-project/), funded by the European Union. Views and opinions expressed are however those of the author(s) only and do not necessarily reflect those of the European Union or the Health and Digital Executive Agency.Neither the European Union nor the granting authority can be held responsible for them.

## 7 Data availability

The data used to parametrize the model are publically available, along with the model code at https://github.com/aiteslya/HIV_MSM_Testing_AB. All analyses performed in this work used synthetic data generated by the agent-based model. The model was implemented in MATLAB R2024b.

## 8 Author Contribution

GR was awarded the funding supporting this research. GR and AT conceptualized the study. AJS, KJJ, MSvdL curated the data. GR, MEK, JAR and AT developed study design. AT revised and maintained the model, ran the simulations and analyzed the data. AT, JAR, JCMH, MSvdL, AvS, AJS, KJJ, MEK, GR interpreted the results. AT and GR wrote the initial draft of the manuscript. AT, JAR, JCMH, MSvdL, AvS, AJS, KJJ, MEK, GR edited and approved the final version of the article.

## 9 Competing interest

Ganna Rozhnova is editorial board member of Communications Medicine.

## Notes

### Competing Interest Statement

The authors have declared no competing interest.

