## Supplementary material for "Comparative evaluation of HIV testing interventions for men who have sex with men in the Netherlands: insights for a low-incidence setting": Methods

December 2025

### 1 Model and parametrization

Below we give a comprehensive description of the model, describe the procedures used to estimate parameter values, and provide the parameters themselves.

We consider the population of men who have sex with men (MSM) in the Netherlands. The estimated size of this population in the Netherlands ranges from 200,000 to 300,000 individuals [13]. For computational tractability we use a population size of 25,000 individuals, which is sufficiently large to yield representative results that can be extrapolated to the national level.

In this study, we assess the impact of the intervention over 15 years. Dynamics of HIV transmission in the population of MSM in the Netherlands are shaped by three processes: demographic processes, sexual network dynamics, and HIV transmission dynamics. Demographic processes include the entry and exit of individuals to and from the population, as well as ageing. Sexual network dynamics are shaped by the formation and dissolution of sexual partnerships that facilitate contacts, facilitating effective contacts where HIV acquisition is possible. Lastly, HIV transmission dynamics include the natural course of HIV infection in an exposed individual, combined with care and prevention measures: diagnosis, ART initiation and support within cascade of care, and a pre-exposure prophylaxis (PrEP) program.

We use a stochastic agent-based model. To capture variability of outcomes in a given scenario, we collect and analyzed ensembles of simulation trajectories. By calibrating the model to the Dutch HIV surveillance data in MSM population, we have selected the 100 best parameter sets. For each scenario, we simulate 20 trajectories per parameter set, using distinct random seeds to introduce variability.

#### 1.1 Simulation detail

At the start of the simulation we initiate model population of 25,000 individuals. The simulation progresses with constant one-week steps. At each step, demographic, sexual network, and HIV transmission events can occur based on the population-state-dependent probability. The simulation run comprises four distinct stages: (a) An initial two-year burn-in stage, during which sexual network dynamics settle to a pseudo-equilibrium state without activating HIV processes; (b) The subsequent 7.5 years dedicated to establishing the epidemiological state of the population, where the infection profile of the population is initialized and evolves governed by demographic processes, sexual network dynamics and HIV transmission dynamics. This stage of the simulation is used to calibrate the model by matching the evolution of HIV dynamics in the population of MSM during the 2017-2023 period; (c) Two year run during 2024–2025 with baseline interventions being active (the parameters of PrEP programme and HIV cascade of care following positive diagnosis remain unchanged from the values used during HIV dynamics calibration period); (d) The final 15-year stage during which intervention scenarios are modeled. The intervention timeline commences on July 1, 2026.

#### 1.2 Demography

The demographic module includes the entry of individuals into the population, exit due to non–HIV-related (background) mortality, and ageing. In the MSM population, HIV transmission occurs primarily through condomless anal intercourse (AI) [3]. Accordingly, we model sexually active men aged 15–74 years, grouped into 10-year age bands: 15–24, 25–34, 35–44, 45-54, 55–64, and 65–74 years. Individuals enter the population according to the distribution of age at sexual debut, based on The European MSM Internet Survey 2017 (EMIS-2017) data for the Netherlands [30] (Figure 1). The responses ranged from 15 to 30 years and greater than 30 years.

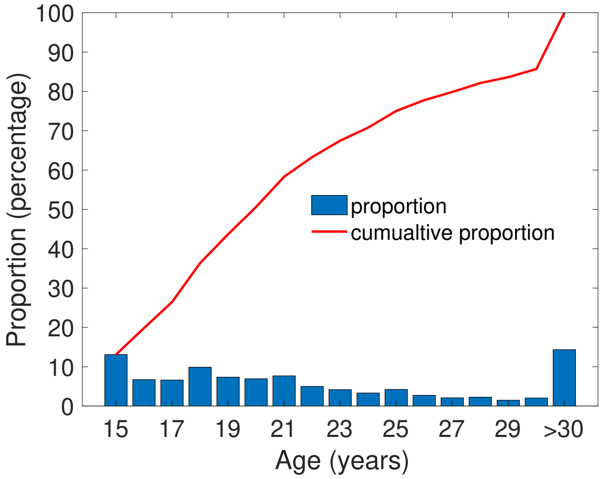


Figure 1: **Distribution of age at sexual debut reported by EMIS-2017 respondents.** Bars represent the proportion of individuals whose sexual debut occurred at each specific age, while the red curve shows the cumulative proportion of respondents who had sexual debut at or before that age.

At model initialization, the age distribution of individuals is set to reflect the demographic equilibrium expected under the distribution of age of sexual debut and current mortality rates in the Netherlands. This distribution is obtained from a corresponding system of ordinary differential equations describing demographic dynamics. For individuals whose age of sexual debut exceeds 30, the exact age is assigned by uniformly sampling within 31–74 age band. Age affects individual characteristics, including the likelihood of forming non-steady partnerships, preferred age for partners (both steady and non-steady), condom use, and background mortality rate. While the model includes individuals who immigrate to the Netherlands, we do not further stratify by ethnicity. Individuals exit the population upon reaching age 75, representing the cessation of sexual activity, or earlier due to age-dependent background mortality [1]. Mortality within each age band follows a geometric process, with age-specific rates derived from WHO life tables for the Netherlands [1]. Individuals who leave the population are not immediately replaced. Age increases continuously over time, with each individual ageing by one year every 52 simulation weeks. Upon moving to a new age band, the corresponding mortality rate is updated. The appearance of new individuals is modelled as a Poisson process with mean rate $\lambda$ per year, calculated from the equilibrium condition of the analogous deterministic model. This ensures a quasi-stable population size throughout the simulation. Parameters related to demographic dynamics are listed in Supplementary Table 1.

| Description (unit) | Value | Source |
|---|---|---|
| Time spent in each age band (years) | 10 | Follows from the definition of age bands |
| Background death rate in age band (year$^{-1}$) | | WHO life tables for the Netherlands [1] |
| 15-24 | $0.3 \times 10^{-3}$ | |
| 25-34 | $0.5 \times 10^{-3}$ | |
| 35-44 | $0.9 \times 10^{-3}$ | |
| 45-54 | $2.4 \times 10^{-3}$ | |
| 55-64 | $7.0 \times 10^{-3}$ | |
| 65-74 | $20.5 \times 10^{-3}$ | |
| Mean rate of entrance of new individuals (individuals year$^{-1}$) | 4,280 | Derived from the deterministic analog model, using 200,000 individuals as an equilibrium population size. |

Table 1: **Demographic processes: summary of model parameters.**

#### 1.3 Sexual network dynamics

To accurately represent HIV transmission among MSM, the model captures the evolving structure of the sexual network and its dependence on age profile of the population. Two types of partnerships are distinguished: steady and non-steady. Following EMIS-2010 [29] and EMIS-2017 [30], a steady partnership refers to a long-term relationship in which partners do not consider themselves single. In contrast, a non-steady partnership includes all other sexual contacts—such as one-time encounters, one-night stands, casual relationships without a defined arrangement, and sex-buddy partnerships. The main distinctions between steady and non-steady partnerships lie in (1) relationship duration, (2) frequency of AI, (3) proportion of condomless AI, and (4) degree of concurrency. Partner selection for both partnership types depends on age and HIV status, reflecting age assortativity and serosorting. Parameters governing sexual network dynamics are summarised in Supplementary Table 2.

##### 1.3.1 Steady partnerships

Analysis of EMIS-2017 data shows that most MSM when asked about existence of a steady partnership reported either having no steady male partner (approximately 55%) or being in a steady partnership with one man (around 42%), while a small fraction (approximately 3%) reported being involved in a steady partnership with more than one man. Accordingly, in the model each individual can either be single or in a steady partnership with one man at any given time. The simulation maintains a proportion of individuals with a steady partner fluctuating around 45%. At each time step, a number of new steady partnerships is formed while a number of others dissolve. The number of new partnerships per unit time depends on the number of single individuals and the steady partnership formation rate, while the number of dissolutions depends on the average partnership duration. All single individuals are assumed to have the same probability of forming a steady partnership, and partnership duration follows a geometric distribution with a fixed mean.

At model initialization, a sufficient number of steady partnerships is formed to achieve a quasi-equilibrium such that 44% of individuals are in steady relationships. For each new pair, the first partner is selected randomly from the pool of single individuals. Their age group and HIV status determine preferences for age and serostatus of potential partners (Supplementary Tables 3 and 5). The second partner is then sampled from individuals matching the preferred age band and HIV status who are not already in a steady partnership. If two selected individuals are not currently involved in a non-steady partnership, a new pair is formed.

As the simulation proceeds, new partnerships are continuously formed and dissolved, keeping the proportion of individuals with a steady partner near equilibrium. Each partnership is assigned a duration at the time of formation, and partnerships are dissolved once their end date coincides with the current simulation time, after which both partners re-enter the pool of single individuals. Both formation and dissolution rates are calibrated to match the proportion of individuals in steady partnerships and the median number of steady partners in the past 12 months reported by EMIS-2017 respondents [30].

The modelling framework underlying partnership formation follows the pair-formation approach introduced by Dietz and Hadeler [4] and later extended by Kretzschmar and colleagues [10, 12, 11]. Analysis of the deterministic analogue model shows that the number of individuals in steady partnerships stabilises around a steady-state value determined by the partnership formation rate and average duration.

##### 1.3.2 Non-steady partnerships

In addition to steady partnerships, individuals can also form non-steady partnerships. Unlike steady relationships, individuals may engage in non-steady partnerships regardless of whether they already have a steady partner. Moreover, individuals may participate in more than one non-steady partnership at a time. Data show strong heterogeneity propensity to form non-steady partnerships: while some individuals report few non-steady partners (0–2) over six months, others report many more (over six in the same period, Figure 2).

Data from the Amsterdam Cohort Studies (ACS) indicate that most non-steady partnerships are brief, typically lasting only a few days. Therefore, the pair-formation framework used for steady partnerships is not well suited for modelling these contacts. Instead, we assign each individual an intrinsic propensity to form non-steady partnerships. Individuals with higher propensity values have a greater chance of being forming a partnership than those with lower values. Analysis of ACS data shows that the rate of non-steady partner acquisition depends on age and on whether an individual currently has a steady partner (Supplementary Figure 2). We calculated the distribution of the number of non-steady partners in the previous six months, stratified by age group and steady-partner status. These distributions were normalised and used to define propensities for forming non-steady partnerships. Propensity values are continuously updated as individuals age or change steady-partner status.

As with steady partnerships, new non-steady partnerships can form and existing ones can dissolve at any point in time. The number of new partnerships formed depends on the population size and the overall formation rate. When a partnership is to be formed, the first partner is selected with probability proportional to individual propensity. The index partner's age and HIV status determine the distributions for partner selection by age and serostatus (Tables 4 and 5), from which the characteristics of the second partner are sampled. The second partner is then chosen from the subset of individuals matching these characteristics, also with probability proportional to their propensity. A new pair is formed if the two selected individuals are not already in a partnership together. The duration of each non-steady partnership is sampled from an exponential distribution. As the simulation progresses, each partnership's scheduled end time is continuously checked against the current simulation time, and the partnership is dissolved when its duration expires.

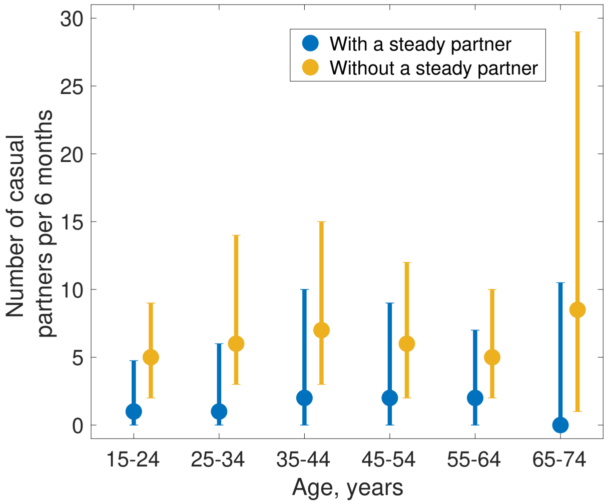


Figure 2: **Distribution of acquisition rate of non-steady partners by age, with and without a steady partner.** Analysis of the rates of acquisition of non-steady partners per 6-month period using data Amsterdam Cohort Studies (ACS) [9]. Large dots denote medians of distribution with whiskers denoting 50-th and 75-th quartiles.

To estimate partnership formation rates we analysed MSM Network Study (NS) data from Amsterdam [8, 14], from which we obtained a mean duration of non-steady partnerships of 2 days. The Amsterdam Cohort Study provides the median number of non-steady partners over the preceding 6 months.

For both steady and non-steady partnerships model calibration employed a Nelder–Mead optimisation (MATLAB's fminsearch) to estimate steady- and non-steady–partnership formation rates and the expected duration of steady partnerships, using parameter values from the equilibrium of the deterministic pair-formation model as initial seeds. Between-partners times and partnership durations are assumed to be exponentially distributed.

| Description (unit) | Value | Source |
|---|---|---|
| Mean duration of a steady partnership (year) | 1.64 | MSM Network Study in Amsterdam [8, 14] |
| Formation rate of steady partnerships ($year^{-1}$) | 0.57 | EMIS-2017 [30] |
| Mean duration of a non-steady partnership (day) | 2.0 | MSM Network Study in Amsterdam [8, 14] |
| Formation rate of non-steady partnerships ($day^{-1}$) | 0.02 | ACS [9] |

Table 2: **Summary of model parameters and transition rates: sexual network dynamics.**

In our previously published work using the same model [18], we validated the model's behavioural dynamics by comparing partner rate-based class transitions with those reported by Basten et al. for HIV-negative MSM, observing close agreement in class stability and the predominance of shifts to adjacent classes [2]. Initialising the population to the empirical distribution of risk categories, the model reproduced a 6-month transition matrix consistent with the cohort data, including a greater likelihood of medium-to-low than medium-to-high transitions.

##### 1.3.3 Age assortativity and serosorting

To obtain age-dependent and sero-dependent mixing patterns in the population, we analyzed data collected in the MSM Network study [8, 14].

To calculate age-mixing matrices for steady and non-steady partnerships we used the method described in [27]. In short, this method allows the estimation of contact frequency between different age groups by employing the maximum likelihood method to estimate the mean number of contacts between individuals in different age bands, while correcting for reciprocity of contacts by modifying the log-likelihood function using weights of relative size of each age band with respect to the total population. Once the rates were calculated, we normalized the contact rates for each age band, to obtain age preference distributions for each age band (Tables 3 and 4).

| | Age of the individual | | | | | |
|---|---|---|---|---|---|---|
| **Age of partner** | 15–24 | 25–34 | 35–44 | 45–54 | 55–64 | 65–74 |
| 15–24 | 0.3724 | 0.1592 | 0.0616 | 0.0392 | 0.0395 | 0.0000 |
| 25–34 | 0.4563 | 0.4866 | 0.3197 | 0.2392 | 0.2057 | 0.1470 |
| 35–44 | 0.1539 | 0.2811 | 0.4555 | 0.4478 | 0.3935 | 0.4526 |
| 45–54 | 0.0174 | 0.0589 | 0.1538 | 0.2465 | 0.2813 | 0.4004 |
| 55–64 | 0.0000 | 0.0105 | 0.0094 | 0.0245 | 0.0700 | 0.0000 |
| 65–74 | 0.0000 | 0.0037 | 0.0000 | 0.0028 | 0.0100 | 0.0000 |

Table 3: **Age-dependent mixing preference for individuals in steady partnerships.**

| | Age of the individual | | | | | |
|---|---|---|---|---|---|---|
| **Age of partner** | 15–24 | 25–34 | 35–44 | 45–54 | 55–64 | 65–74 |
| 15–24 | 0.4002 | 0.1633 | 0.0662 | 0.0377 | 0.0497 | 0.1036 |
| 25–34 | 0.4184 | 0.5101 | 0.3670 | 0.2409 | 0.1972 | 0.1363 |
| 35–44 | 0.1306 | 0.2753 | 0.4489 | 0.4705 | 0.4444 | 0.3216 |
| 45–54 | 0.0421 | 0.0428 | 0.1082 | 0.2253 | 0.2499 | 0.3343 |
| 55–64 | 0.0086 | 0.0066 | 0.0097 | 0.0242 | 0.0588 | 0.1042 |
| 65–74 | 0.0000 | 0.0019 | 0.0000 | 0.0014 | 0.0000 | 0.0000 |

Table 4: **Age-dependent mixing preference for individuals in non-steady partnerships.** Each column is normalized to add up to 1.

To identify tendencies of MSM to serosort, we analyzed the MSM Network Study data. In this study, the respondents reported the knowledge or perception of their own HIV status, as well as the knowledge or perception of the HIV status of their partner. Based on the data, and adjusting for the prevalence of individuals with diagnosed HIV infection in the population of MSM, we have calculated matrices describing the probability distribution of sexual mixing for steady and non-steady partnerships (Supplementary Table 5). We adjusted the serosorting matrix in formation of non-steady partnerships, we have taken into the account the proportion of non-steady partnerships with an annonymous partner. In effect, the serosorting delineates the population into two subgroups: those who have received an HIV diagnosis and those who have not.

| | **Steady partnerships** | | **Non-steady partnerships** | |
|---|---|---|---|---|
| | **No HIV diagnosis** | **HIV diagnosed** | **No HIV diagnosis** | **HIV diagnosed** |
| **No HIV diagnosis** | 0.981 | 0.254 | 0.982 | 0.688 |
| **HIV diagnosed** | 0.019 | 0.746 | 0.018 | 0.312 |

Table 5: **Serosorting patterns distributed by the type of the partnership.** Each column is normalized to add up to 1.

#### 1.4 HIV dynamics

This section describes the model of the natural history of HIV transmission in MSM and care and prevention interventions in place in the Netherlands in 2023.

##### 1.4.1 Natural progression of infection.

The model of HIV natural history in MSM, excluding HIV care and prevention services (testing, ART initiation, and PrEP), follows a standard framework. In this framework, susceptible individuals are exposed through condomless anal intercourse (AI) with an infectious partner, potentially leading to infection. Individuals who newly acquired HIV enter a brief early infection stage, followed by a prolonged chronic stage, and then progress to AIDS, which we divide into early and late sub-stages. Individuals with HIV may transmit in all stages after acquisition, and, therefore, the exposure risk to susceptible individuals increases with the prevalence of people living with HIV within their local sexual network.

The early stage is short (mean duration 89 days [6]) and marked by elevated viral load and, consequently, a higher transmission probability. It is followed by a chronic stage (mean duration 8.31 years [12]) with lower relatively stable viral load and few clinical manifestations, after which individuals enter early AIDS, characterised by opportunistic infections and AIDS-related malignancies (mean duration 1.18 years [12]), and then late AIDS with rapidly rising viral load and multisystem disease. Both AIDS sub-stages carry additional mortality, with mean survival period after AIDS onset of 3 years [12]. To avoid unrealistically short early HIV infection, we split it into three Fiebig-based sub-stages (1; 2–3; 4–5) with mean durations of 5, 9, and 75 days, respectively [6]. Times in early infection and AIDS are modelled as exponential; the chronic duration follows an Erlang distribution (shape parameter 80) to limit variance.

Stage-specific viral load profiles determine per-act transmission potential. Using literature estimates [12], we set the chronic stage as baseline, with a 26-fold higher transmission potential in early HIV infection and a 6-fold increase in early AIDS. While individuals in the late AIDS stage have a still higher potential to transmit, due to severe morbidity they are assumed not to be sexually active. The per-act transmission probability in chronic infection is obtained by calibrating the model to annual new HIV infections and diagnoses.

The daily transmission probability combines the AI rate, the probability of condom use per AI, and the per-act transmission probability for condomless AI. From the MSM Sexual Network survey [8, 14], we estimate a mean AI rate of 0.33 contacts per day in steady partnerships and 0.11 per day in non-steady partnerships. To ensure at least one AI event per non-steady partnership, each such partnership is initiated with an AI act.

To estimate the probability of condom use, we analysed the proportion of steady and non-steady partners with whom respondents of EMIS-2017 data set [31] reported having condomless AI. Analysis of the data set yielded that condom use during AI differed across different types of partnerships and ages of individuals (Supplementary Table 6). We reconcile condom use by taking the highest value between the two partners.

| | **Age of the individual** | | | | | |
|---|---|---|---|---|---|---|
| **Partnership Type** | 15–24 | 25–34 | 35–44 | 45–54 | 55–64 | 65–74 |
| **Steady** | 0.29 | 0.22 | 0.28 | 0.28 | 0.28 | 0.27 |
| **Non-Steady** | 0.63 | 0.65 | 0.58 | 0.54 | 0.55 | 0.48 |

Table 6: **Condom use probability in steady and non-steady partnerships distributed by age.**

##### 1.4.2 Care and prevention programmes

**Diagnosis and cascade of care.** Individuals with HIV can be diagnosed, linked to care and initiate treatment with the goal of achieving an undetectable viral load.

Testing rates depend on the number of non-steady partners within the previous 6 months and PrEP use. The model of testing process and parametrization is described in detail in Subsection 2.1 Model of Section 2 Methods in the main text.

Following diagnosis, individuals initiate ART and subsequently achieve viral load suppression (less than 200 copies/mL). Both the diagnosis-to-ART and ART-to-suppression intervals are modelled as exponentially distributed. Final parameter values were obtained by calibrating to cascade-of-care trends among MSM in the Netherlands around the index values. The mean time form diagnosis to ART was taken to be 2 months; shorter than 2010 practice but longer than current practice (Chapter 1, [26]). The average time between ART initiation to achieving viral suppression was guided by the findings of Dijkstra et al. [5] (median 55 days, CI 31–72). During the first year following the diagnosis, diagnosed individuals reduce contact rates in both partnership types by a factor of 1.3 [7].

Finally, a fraction of individuals can drop out of treatment or may experience treatment failure, as the result, their the viral load is no longer suppressed. In both cases, we model that individuals move to the stage corresponding to individuals with chronic HIV infection who were diagnosed. We determined the ART failure/dropout rate by initially selecting an interval based on data showing the proportion of individuals who had not achieved viral suppression two years after initiating ART (see Figure 2.10D, Chapter 2 in [26]), and then refined this interval through the calibration process.

**PrEP.** In the Netherlands, a national PrEP program prioritizing people at risk of acquiring HIV, of whom MSM are a substantial portion, started in 2019. Before this time, PrEP uptake was low and those who used it either procured it from abroad or received it via the AMPrEP demonstration study which ran in Amsterdam, the Netherlands, from 2015 to 2020. In addition to the national PrEP program, PrEP may be obtained via general practitioners. The number of MSM currently enrolled in the PrEP program is capped at 8,500 with an additional 1,000 procuring PrEP from abroad and general practitioners; thus, if we assume a population of 200,000 individuals, i.e., an estimated 5.0% uses PrEP. We modeled PrEP uptake as starting with no users at the end of 2018, PrEP use starting in 2019, and gradually growing towards a level of 5.0% by January 2022, after which time the the percentage of PrEP users fluctuates around this value. While PrEP can be taken either as a daily preparation or timed around the anticipated time of sexual activity, for simplicity we modeled the use as continuous.

Population which uses PrEP changes dynamically, such that at each simulation step, a number of individuals fulfilling the criteria for PrEP program enrolment start using PrEP. Similarly, individuals who are currently using PrEP can cease doing so. Both enrolments to and leaving the program are modeled using exponential waiting times. Since the goal of the PrEP program is to protect individuals most at risk of HIV acquisition, we approximated the PrEP eligibility criteria using the number of non-steady partners within the last six months. Individuals whose number of partners exceeded a threshold, were eligible to enrol. We modelled an average PrEP enrollment as an exponential process with mean of 2 years [28]. When individuals enrolled to the programme, they were assigned a termination time at which point the PrEP use was discontinued. PrEP could also be discontinued as a result of leaving the population or the eligibility criteria being no longer applicable.

According to the available evidence, individuals with perfect PrEP adherence experience an 86% reduction in the per-act probability of HIV acquisition during condomless AI [16]. In the model, we assume perfect adherence for all PrEP users; accordingly, their per-act HIV acquisition probability during condomless AI is reduced by 86%.

**Importation of new infections** The model includes the acquisition of HIV infection from outside of the local network. When an individual enters the population, their HIV status may be susceptible, HIV-positive without diagnosis, and HIV-positive with diagnosis. The entrance rate of individuals with undiagnosed HIV is a constant parameter whose value is determined through calibration process. The entrance rate of diagnosed individuals is based on the available data collected from yearly Stichting HIV monitoring (SHM) reports [20, 21, 22, 23, 24, 25, 26], years 2017-2022. To extrapolate the rates of arrival of individuals starting 2023 and onwards, we have fitted a linear function (Supplementary Figure 3**A**). The temporary reduction and subsequent rebound in arrival rates during and shortly after COVID-19 pandemic years were treated as representative dynamics and retained within the calibration dataset.

Finally, we have analyzed the proportion of individuals with a suppressed viral load among these that enter the Netherlands with diagnosed HIV infection - it remained constant throughout 2014–2022 (Supplementary Figure 3**B**) and was approximately equal to 95%. Thus, we fixed it as a constant in the simulation.

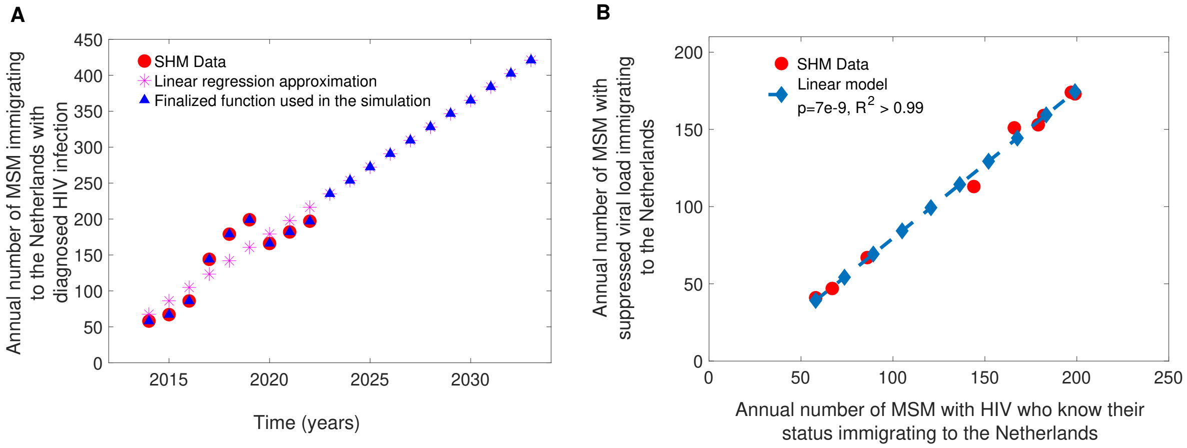


Figure 3: **Entrance rate of individuals with diagnosed HIV.** (a) Annual entrance rates of individuals with diagnosed HIV. Red dots indicate SHM data, magenta stars - linear regression approximation based on the data, blue triangles - composite curve used in the simulations. (b) Analysis of changes in the proportion of individuals with suppressed viral load among those who enter the Netherlands with diagnosed HIV infection.

### 2 Initialization and simulation run

At the start of simulation, we initialise the population's age distribution to match the 2014 age structure of the general male population in the Netherlands [17]. Subsequently, we initiate a number of steady partnerships so that the proportion of individuals in steady partnerships aligns with EMIS-2017 [30], see Subsubsection Steady partnerships, Subsection Sexual network dynamics, section Model and parametrization. We then assign the propensity of forming non-steady partnerships, taking into account each individual's age and whether they currently have a steady partner, see Subsubsection Non-steady partnerships, Subsection Sexual network dynamics, section Model and parametrization.

Once the initialization is complete, the model runs for two years, simulating only demographic and sexual-network processes, to reach a (pseudo)-equilibrium state in sexual-network dynamics. As HIV transmission is not yet active during these first two years of network calibration, serosorting does not influence partnership formation. Once the sexual network is calibrated, the HIV epidemic calibration begins by initialising the epidemic state to match 2016 statistics as captured by SHM data, namely, the total number of MSM living with HIV, the percentage diagnosed among all estimated MSM living with HIV, the percentage on ART among diagnosed MSM, and the percentage of MSM with suppressed viral load among those on ART [20]. For each simulation trajectory, the total number living with HIV is uniformly sampled within the SHM 2016 confidence interval. The specific proportions for diagnosis and the cascade of care are fixed to SHM's point estimates for 2016. The model then proceeds to simulate demographic processes, sexual-network dynamics, and HIV transmission. By the end of simulated 2019, the PrEP programme activates. At the conclusion of the calibration period, which runs until the end of 2023, we allow the simulation to continue for two additional years. When the model time reaches the end of 2025, the main simulation phase begins. Depending on the scenario, this phase may include increased testing in selected subgroups or no further interventions beyond baseline testing rates, the existing care cascade, and the PrEP programme as configured by the end of 2025.

### 3 Calibration and validation

To calibrate the model, we identified a feasible parameter space and sampled it using Latin Hyper Cube Sampling [15] using 4,992 parameter sets. For each set, we created an ensemble of 20 stochastic trajectories and summarized model outputs which we used to match the data with medians. The primary calibration targets were annual numbers of new HIV infections, annual numbers of new HIV diagnoses, annual distribution of stage of infection at the time of the diagnosis as well as the cascade of care in MSM population with respect to the last two steps (proportion of diagnosed MSM who use ART and proportion of MSM on ART who achieved viral load suppression). We matched to the data between 2017-2023, 7 time points per target, leading to matching in order magnitude as well as in the overall trend. We obtained the data estimates from yearly reports published by SHM. To match stage of infection at the time of the diagnosis we used metrics used by SHM, and subsequently delineated the age of infection as follows: 0-6 months, 6-12 months, and greater than 12 months.

We assessed the goodness of fit using the following metric:

$$d\left(v^{\mathrm{t}}, v^{\mathrm{s}}\right) = \max_i \frac{|v_i^{\mathrm{t}} - v_i^{\mathrm{s}}|}{v_i^{\mathrm{t}}} \ , \tag{1}$$

where $v^{\mathrm{t}}$ denotes a target statistic, $v^{\mathrm{s}}$ denotes the respective simulation output (median collected across 20 trajectories), and $i$ takes value between 2017 and 2023, enumerating the years. Observe that this metric is non-dimensionalized, facilitating the goodness of fit assessment across statistics with different dimensions. We selected parameter sets which resulted in the sum of distances between model outputs and the target statistics being below a threshold $\delta$, chosen to be sufficiently small so that 100 parameter sets qualified.

In Table 7 we give the intervals on which parameter values were sampled for calibration.

| Description | Interval (unit) |
|---|---|
| Probability of transmission per CAI | $2.8 - 5.3 \times 10^{-3}$ |
| Incoming rate of HIV infections through immigration | $26 - 48$ (per year) |
| Mean ART initiation rate | $1.8 - 3.2$ (per year) |
| Viral load suppression rate | $1.6 - 2.9$ (per year) |
| ART failure/dropout rate | $3.9 - 7.2 \times 10^{-2}$ (per year) |
| Proportion of individuals in the risk group 1 testing with the second lowest testing rate | 0.36 - 0.66 |
| Proportion of individuals in the risk group 2 testing with the second lowest testing rate | 0.19 - 0.35 |
| Proportion of individuals in the risk group 3 testing with the second lowest testing rate | 0.11 - 0.20 |
| Proportion of individuals in the risk group 4 testing with the second lowest testing rate | 0.18 - 0.34 |
| Threshold value of number non-steady partners in the last 6 months to be eligible to enrol into PrEP programme | 14 - 27 |
| Highest testing rate | 1.2 - 2.3 (per year) |
| Second highest testing rate | 0.37 - 0.69 (per year) |

Table 7: **Sampling intervals for parameters whose distribution was obtained via calibration process.**

In Figure 4 and Table 8 we provide the description of the distribution and means of the parameters which were through calibration process.

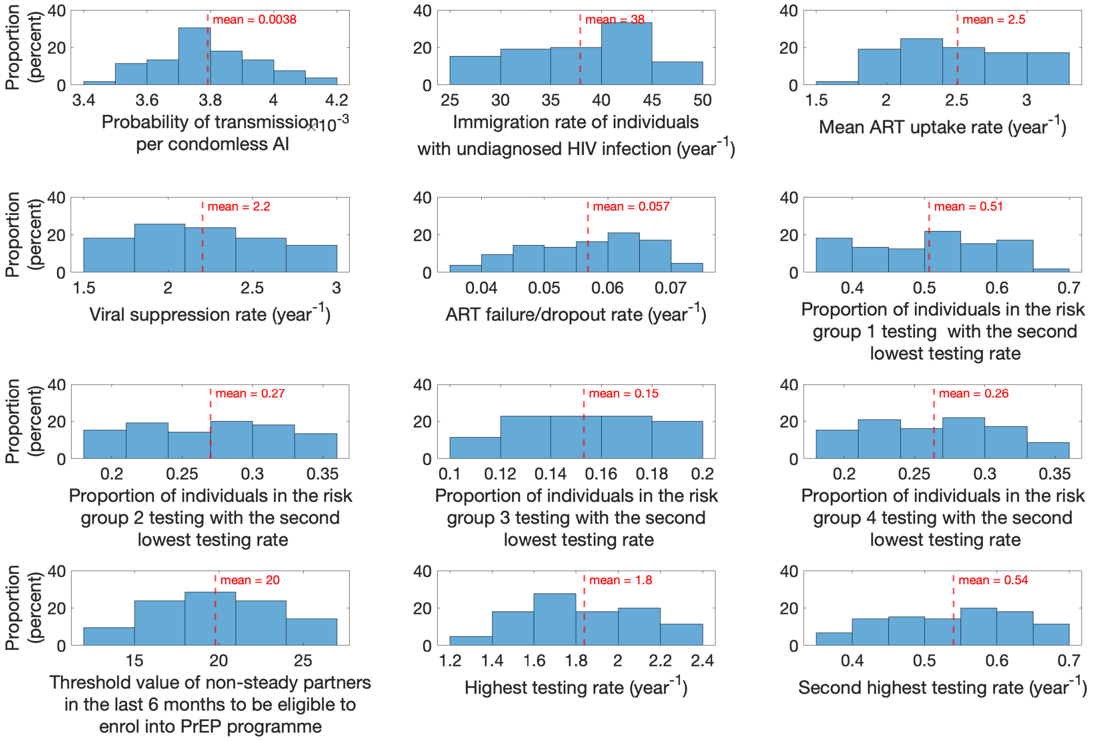


Figure 4: **Parameter distributions used in the simulation.** The distributions were obtained through the calibration process. Dashed red lines indicate mean values.

| Description | Mean (unit) |
|---|---|
| Probability of transmission per CAI | $3.8 \times 10^{-3}$ |
| Incoming rate of HIV infections through immigration | 38 (per year) |
| Mean ART initiation rate | 2.5 (per year) |
| Viral load suppression rate | 2.2 (per year) |
| ART failure/dropout rate | $5.7 \times 10^{-2}$ (per year) |
| Proportion of individuals in the risk group 1 testing with the second lowest testing rate | 0.51 |
| Proportion of individuals in the risk group 2 testing with the second lowest testing rate | 0.27 |
| Proportion of individuals in the risk group 3 testing with the second lowest testing rate | 0.15 |
| Proportion of individuals in the risk group 4 testing with the second lowest testing rate | 0.26 |
| Threshold value of number non-steady partners in the last 6 months to be eligible to enrol into PrEP programme | 20 |
| Highest testing rate | 1.8 (per year) |
| Second highest testing rate | 0.54 (per year) |

Table 8: **Mean values for parameters whose distribution was obtained via calibration process.**

The fit of the model to primary targets is shown on Figure 5. Since these targets were exact measurements, the respective depiction is given without uncertainty intervals.

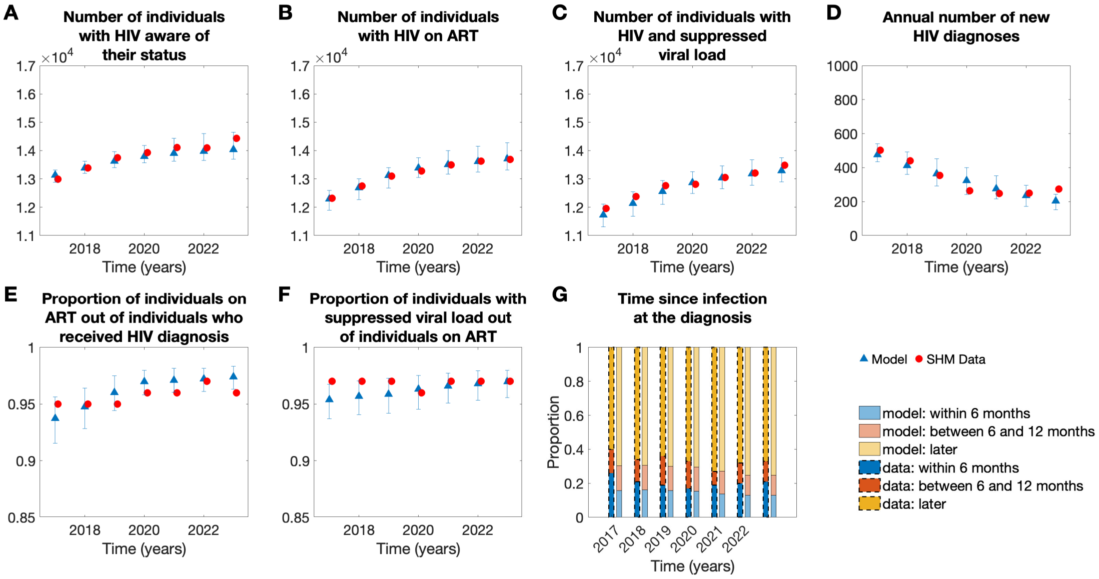


Figure 5: **Calibration of the model** The calibration statistics were time series of **A** Total number of MSM who have received positive HIV diagnosis, **B** Total number of MSM using ART, **C** Total number of MSM with HIV infection and a suppressed viral load, **D** Annual number of new HIV diagnoses in MSM, **E** Proportion of individuals on ART out of MSM who received positive HIV diagnosis (second pillar of cascade of HIV care), **F** Proportion of individuals with suppressed viral load out of MSM using ART (third pillar of cascade of HIV care), **G** Annual distribution of time between HIV acquisition and the day of diagnosis over 2017–2023.

The validation of the model outputs against estimates published by SHM [19, 20, 21, 22, 23, 24, 25, 26] is shown on Figure 6.

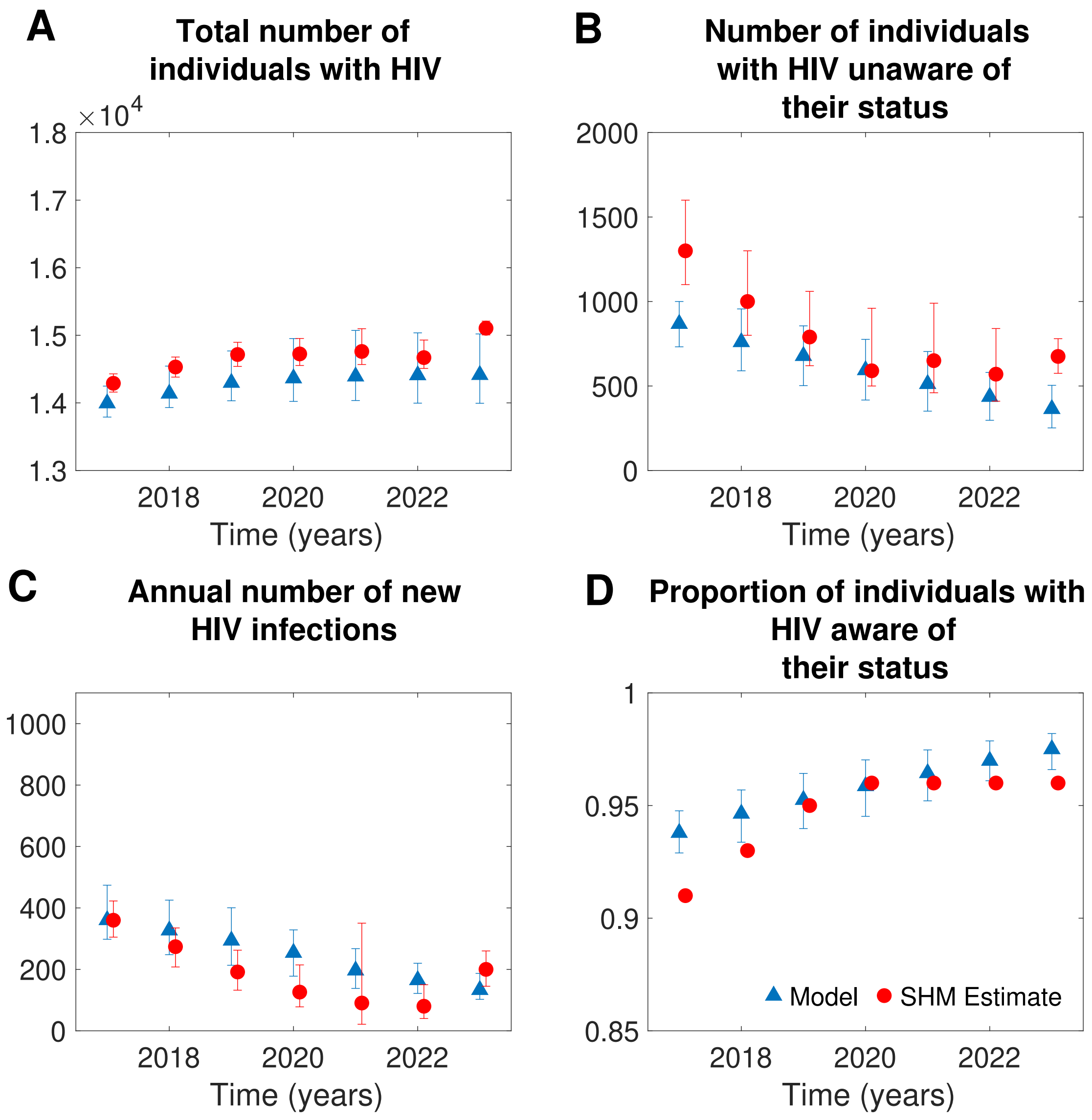


Figure 6: **Validation of the model** The validation statistics were time series of **A** Total number of MSM with HIV infection, **B** Total number of MSM with HIV infection who were not diagnosed yet, **C** Annual number of new HIV infections, and **D** Proportion of MSM with HIV who received positive HIV diagnosis (first pillar of cascade of HIV care) over 2017–2023
