## Additional analyses for "Comparative evaluation of HIV testing interventions for men who have sex with men in the Netherlands: insights for a low-incidence setting"

December 2025

### 1 Time series of baseline dynamics in 2026–2040

In the baseline scenario, with only currently active interventions continuing with the parameters of the ongoing HIV care and prevention programmes remain unchanged from the values used in the calibration period, with an exception of PrEP programme testing frequency which is equal to once in 6 month starting January 2025, the annual incidence of new HIV infections is projected to further decline throughout 2024–2040 (Figure 1).

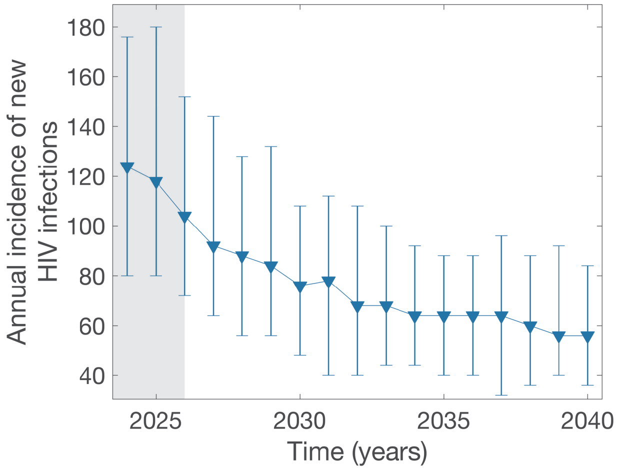


Figure 1: **Annual incidence of new HIV infections in the baseline scenario.** Triangles denote median estimate, with whiskers denoting 95 IR. White area denotes time horizon when intervention is deployed. This projection does not include HIV infections acquired outside the Netherlands.

### 2 Full set of outcomes for the scenarios presented in the main text

In Table 1 we present full range of outputs obtained during the simulations. The results are given in the form of medians across parameter sets and 95% QI.

| Scenario | Measurement | | |
|---|---|---|---|
| | **Cumulative HIV infections averted** **M (95% QI)** | **Additional HIV tests per infection averted** **M (95% QI)** | **Annual additional tests per 100,000 individuals** **M (95% QI)** |
| **1.1** | 16 (-172 – 220) | 564.8 ( – )* | 301.2 ( – )* |
| **1.2** | 46 (-144 – 324) | 491.1 (–) | 753.1 ( – )* |
| **1.3** | 94 (-128 – 328) | 480.7 (–) | 1,506.2 ( – )* |
| **1.4** | 92 (-60 – 344) | 736.7 (–) | 2,259.3 ( – )* |
| **2.1** | 0 (0 – 96) | – ( – ) | 28.5 (14.8 – 54.9) |
| **2.2** | 54 (-144 – 328) | 530.7 (416 – 662.7) | 955.2 (749.5 – 1,192.8) |
| **2.3** | 256 (68 – 464) | 1,122.0 (1,052.0 – 1,206.1) | 9,574 (8,977.3 – 10,292) |
| **2.4** | 0 (-36 – 156) | – ( – ) | 64.1 (35.1 – 179.7) |
| **2.5** | 92 (-72 – 352) | 630.0 (529.0 – 768.9) | 1,932 (1,622.1 – 2,358) |
| **2.6** | 340 (132 – 592) | 1,626.9 (1,526.4 – 1,736.0) | 18,438 (17,299 – 19,674) |
| **3.1** | 6 (-104 – 224) | 1,118 (402.0 – 2,094.7) | 223.6 (80.4 – 418.9) |
| **3.2** | 200 (16 – 424) | 931.3 (814.6 – 1,089.4) | 6,208.7 (5,430.4 – 7,262.4) |
| **3.3** | 508 (292 – 900) | 3,224.3 (3,023.3 – 3,427.9) | 54,598 (51,195 – 58,046) |
| **4.1** | 106 (-116 – 388) | 479.6 (417.7 – 539.5) | 1,694.5 (1,475.9 – 1,906.2) |
| **4.2** | 294 (84 – 528) | 1,059.6 (996.0 – 1,120.3) | 10,384 (9,760.8 – 10,979) |
| **4.3** | 126 (-80 – 364) | 638.9 (562.6 – 739.4) | 2,683.4 (2,363.0 – 3,105.3) |
| **4.4** | 380 (148 – 672) | 1,517.5 (1,431.2 – 1,606.6) | 19,221 (18,129 – 20,351) |
| **4.5** | 154 (-60 – 376) | 475.8 (434.9 – 522.1) | 2,442.4 (2,231.4 – 2,680.2) |
| **4.6** | 308 (156 – 628) | 1,078.7 (1,022.2 – 1,150.8) | 11,075 (10,494 – 11,815) |
| **4.7** | 182 (-32 – 388) | 564.5 (515.4 – 632.0) | 3,424.8 (3,126.7 – 3.834.2) |
| **4.8** | 390 (180 – 708) | 1,532.1 (1,451.4 – 1,626.5) | 19,918 (18,868 – 21,144) |
| **5.1** | 222 (28 – 556) | 940.9 (821.6 – 1,064.5) | 6,962.4 (6,080.0 – 7,877.1) |
| **5.2** | 536 (312 – 792) | 3,079.0 (2,844,6 – 3,255.3) | 55,012 (50,824 – 58,160) |
| **5.3** | 264 (60 – 504) | 878.9 (779.3 – 982.9) | 7,734.0 (6,858.0 – 8,649.6) |
| **5.4** | 534 (308 – 884) | 3,064.8 (2,774.4 – 3,262.2) | 54,553 (49,384 – 58,067) |

Table 1: **Summary of projected dynamics under for intervention scenario.** All quantities are calculated relative to the baseline scenario where intervention parameters are fixed to the values obtained through the calibration process.

#### 2.1 Time series analysis

##### 2.1.1 Annual incidence of new HIV infections

In Figure 2 we show projected time series for the annual number of new HIV infections across different scenarios. The panels on the figures are grouping the series by the type of the intervention and plot them against the time series for the baseline scenario. The exact definitions of interventions can be referred to in Subsection Scenarios, Section Methods in the main text. For all interventions which include the resident MSM as one of the target groups (Interventions 2, 3, 4 and 5) there is a pronounced decrease in projected number of new HIV infections 2 years into the intervention rollout, whereupon intervention infection incidence curves settle into a plateau which is sustained for the remainder of the intervention.

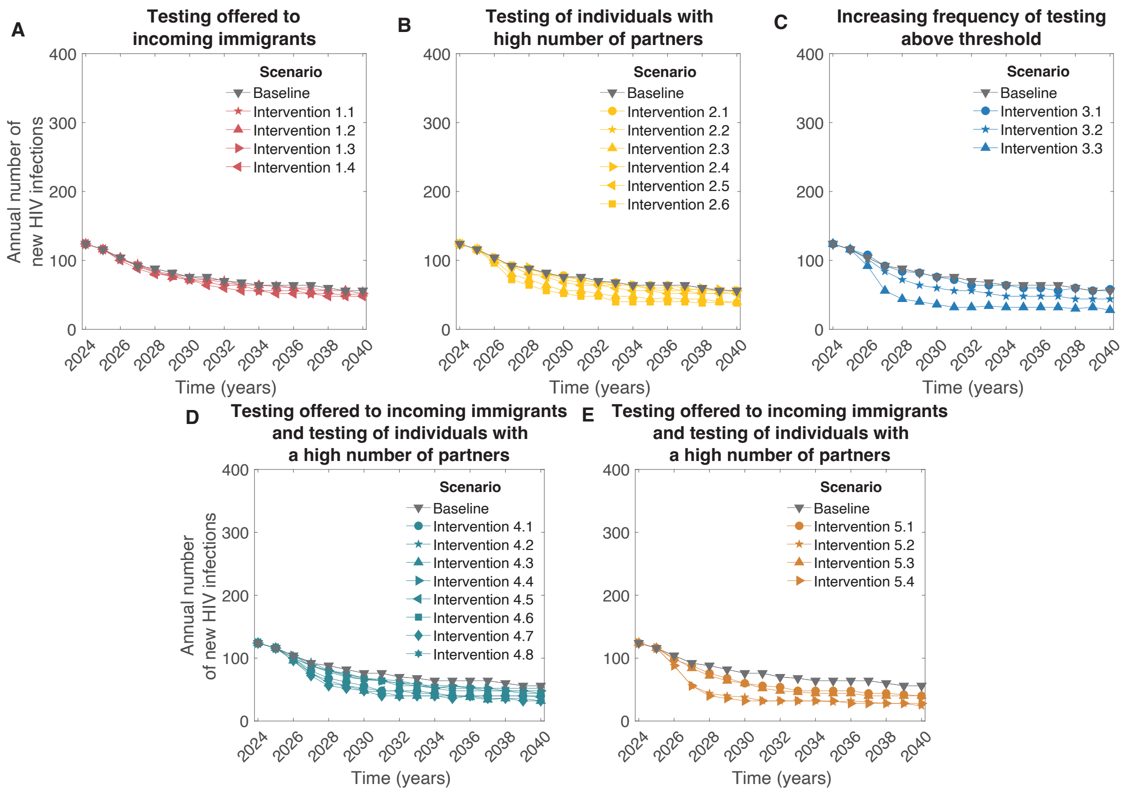


Figure 2: **Annual number of new HIV infections.** **A** One-time testing of individuals immigrating to the Netherlands. **B** Increased testing of individuals with number of non-steady partners in the last 6 months exceeding threshold. **C** Increasing probability to test within 1 year. **D** One-time testing of individuals immigrating to the Netherlands combined with increased testing of individuals with number of non-steady partners in the last 6 months exceeding threshold. **E** One-time testing of individuals immigrating to the Netherlands combined with increasing probability to test within 1 year. On all panels gray triangles denote baseline scenario. The exact definitions of interventions can be referred to in Subsection Scenarios, Section Methods in the main text.

##### 2.1.2 Annual incidence of new HIV diagnoses

The cumulative number of HIV diagnoses is shaped by several factors: incidence of new HIV infections, population size of individuals not yet aware of having HIV and frequency of testing. To isolate these effects, we look at the time series of the annual number of new HIV diagnoses (Figure 3). We observe that for the interventions where a notable decrease in the cumulative number of new HIV infections is expected, the diagnosis time series are characterized by a sharp increase relative to the baseline level in the first years following the intervention start, and then by a fall below the baseline level in the subsequent years, reflecting decreased annual rate of appearance of new HIV infections. The sharper initial increase reflects a stronger subsequent reduction in HIV incidence, so a rise in diagnoses shortly after implementation can serve as an indicator of intervention success.

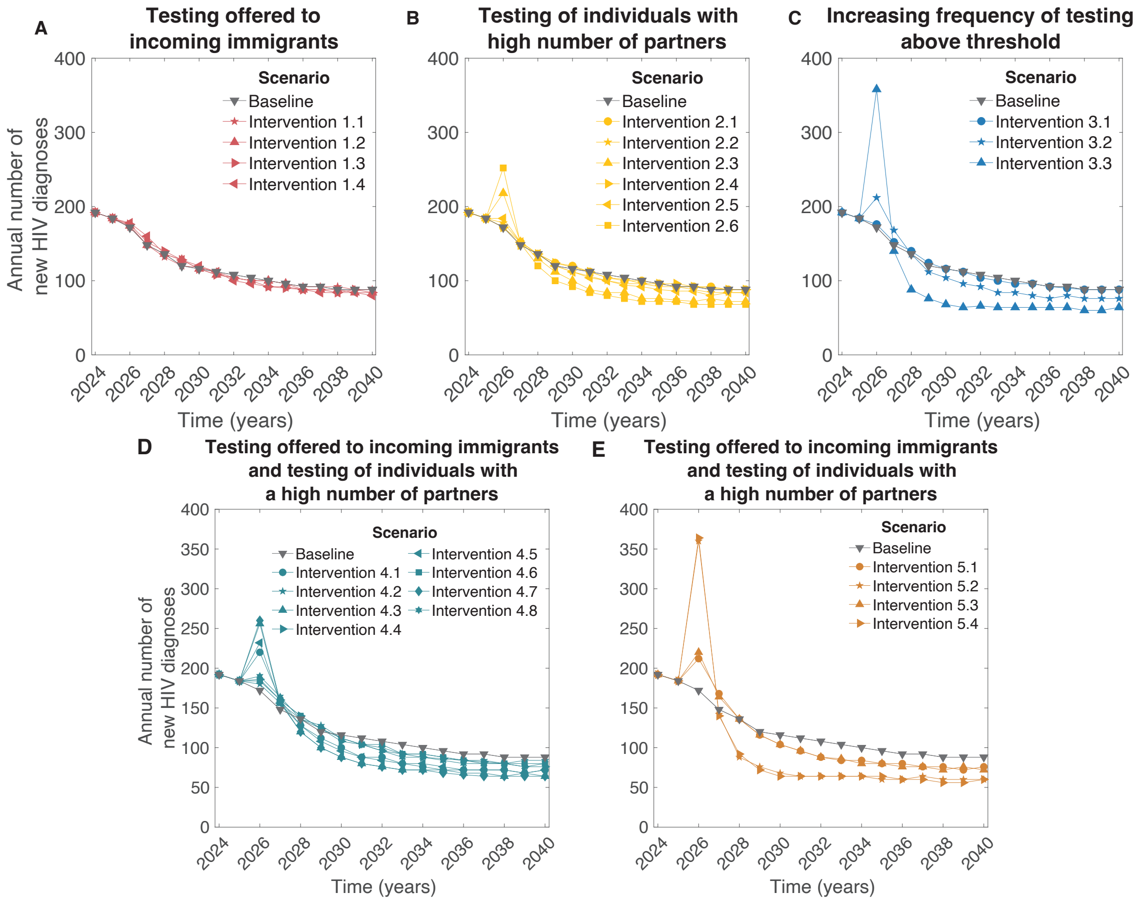


Figure 3: **Annual number of new HIV diagnoses.** **A** Offering a one-time HIV test to incoming immigrants. **B** Increased testing of individuals with number of non-steady partners in the last 6 months exceeding threshold. **C** Increasing probability to test within 1 year. **D** One-time testing of individuals immigrating to the Netherlands combined with increased testing of individuals with number of non-steady partners in the last 6 months exceeding threshold. **E** One-time testing of individuals immigrating to the Netherlands combined with increasing probability to test within 1 year. On all panels gray triangles denote baseline scenario. The exact definitions of interventions can be referred to in Subsection Scenarios, Section Methods in the main text.

### References
